# COVID-19 Quarantine Reveals Grade-specific Behavioral Modification of Myopia: One-Million Chinese Schoolchildren Study

**DOI:** 10.1101/2020.11.15.20231936

**Authors:** Liangde Xu, Yunlong Ma, Jian Yuan, Yaru Zhang, Hong Wang, Guosi Zhang, Changsheng Tu, Xiaoyan Lu, Jing Li, Yichun Xiong, Fukun Chen, Xinting Liu, Zhengbo Xue, Meng Zhou, Wen-Qing Li, Nan Wu, Hao Chen, Jiangfan Chen, Fan Lu, Jianzhong Su, Jia Qu, on behalf of Myopic Epidemiology and Intervention Study

## Abstract

**Background:** High prevalence of myopia of adolescent has been a global public health concern. Their risk factors and effective prevention methods for myopia across schoolchildren developmental stages are critically needed but remain uncertain due to the difficulty in implementing intervention measurements under normal life situation. We aimed to study the impact of the COVID-19 quarantine on myopia development among over one-million schoolchildren.

**Methods:** We designed the ongoing longitudinal project of Myopic Epidemiology and Intervention Study (MEIS) to biannually examine myopia among millions of schoolchildren for ten years in Wenzhou City, Zhejiang Province, China. In the present study, we performed three examinations of myopia in 1,305 elementary and high schools for schoolchildren in June 2019, December 2019 and June 2020. We used the normal period (June-December 2019) and COVID-19 quarantine period (January-June 2020) for comparisons. Myopia was defined as an uncorrected visual acuity of 20/25 or less and a spherical equivalent refraction (SER) of -0.5 diopters (D) or less. High myopia was defined as an SER of -6.0 D or less.

**Findings:** In June 2019, 1,001,749 students aged 7-18 were eligible for examinations. In the 6-month and 12-month follow-up studies, there were 813,755 eligible students (81.2%) and 768,492 eligible students (76.7%), respectively. Among all students, we found that half-year myopia progression increased approximate 1.5 times from -0.263 D (95% CI, -0.262 to -0.264) during normal period to -0.39 D (95% CI, -0.389 to -0.391) during COVID-19 quarantine (P < 0.001). Multivariate Cox regression analysis identified grade rather than age was significantly associated with myopia (Hazard ratio [HR]: 1.10, 95% CI, 1.08 to 1.13; P < 0.001) and high myopia (HR: 1.40, 95% CI, 1.35 to 1.46; P < 0.001) after adjustment for other factors. The prevalence, progression, and incidence of myopia and high myopia could be categorized into two grade groups: I (grades 1-6) and II (grades 7-12). Specifically, COVID-19 quarantine for 6 months sufficiently increased risk of developing myopia (OR: 1.36, 95% CI, 1.33 to 1.40) or high myopia (OR: 1.30, 95% CI, 1.22 to 1.39) in Grade Group I, but decreased risk of developing myopia (OR: 0.45, 95% CI, 0.43 to 0.48) or high myopia (OR: 0.57, 95% CI, 0.54 to 0.59) in Grade Group II.

**Interpretation:** The finding that behavioral modifications for six months during COVID-19 quarantine sufficiently and grade-specifically modify myopia development offers the largest human behavioral intervention data at the one million scale to identify the grade-specific causal factors and effective prevention methods for guiding the formulation of myopia prevention and control policies.

**Funding:** Key Program of National Natural Science Foundation of China; the National Natural Science Foundation of China; Scientific Research Foundation for Talents of Wenzhou Medical University; Key Research and Development Program of Zhejiang Province.

**Research in context:** *Evidence before this study:* Myopia is the most-common refractive error worldwide. Myopia with younger onset may result in developing high myopia, which is associated with sight-threatening ocular diseases such as maculopathy, retinal detachment, opticneuropathy, glaucoma, retinal atrophy, choroidal neovascularization. In light of the increasing prevalence of myopia and high myopia has been a global public health concern, the impact of COVID-19 lockdown on myopia development has gained substantial attention. We searched PubMed, Google Scholar, and MEDLINE databases for original articles reported between database inception and November 10, 2020, using the following search terms: (coronavirus OR COVID* OR SARS-COV-2 OR lockdown OR quarantine) AND (myopia OR short-sightedness OR refractive error). To date, there was no original study reported to uncover the influence of COVID-19 quarantine on myopia progression.

*Added value of this study:* This study provides the largest longitudinal intervention data on myopia progression in Chinese schoolchildren covering all grades of schoolchildren at one-million scale. COVID-19 quarantine model uncovers that behavioral modifications for six months may lead to significant increase of overall prevalence of myopia associated with their increased screen times and decreased outdoor activity times. Importantly, their effects on developing myopia or high myopia of students are grade-dependent, which were risk factors for elementary schools period but protective factors for high schools period partly due to reduced school education burden.

*Implications of all the available evidence:* This one-million schoolchildren myopia survey offers evidence that six months behavioral modifications sufficiently and grade-specifically change the progression of myopia and high myopia. In view of the increased use of electronic devices is an unavoidable trend, effective myopia prevention strategy according to grade among students is urgently needed. Since COVID-19 outbreak is still ongoing and spreading, international collaborate efforts are warranted to uncover the influence of COVID-19 on myopia progression to further substantiate these findings.

## Introduction

Myopia is the most-common cause of visual impairment worldwide ^1^. Many studies have reported the prevalence of myopia among schoolchildren, which ranges from 20% in early school-aged children to as high as 80% among 17-year-olds, in China ^2-6^ and other East and Southeast Asia countries such as Singapore and Japan ^7-9^. The incidence of myopia also tends to be gradually increasing in populations of European and Middle Eastern ancestry ^10, 11^. The latest World Health Organization (WHO) World Report on Vision (issued Oct 8, 2019) ^12^ indicated there were at least 312 million people <19 years of age with myopia in 2015, and myopia was estimated to affect 2.6 billion people of all ages worldwide by 2020, and predicted to affect approximately 5 billion people by 2050, including 938 million people with high myopia^1^. In addition, myopia with younger onset may progress to high myopia ^13^, which are at a substantially increased risk of sight-threatening complications such as myopic maculopathy, glaucoma, and retinal detachment. The increasing prevalence of myopia is a global public health concern^14, 15^, promoting many countries including China ^16^ to implement nationwide policies for preventing myopia development.

The progression of myopia can be influenced by genetic and environmental components^17^. Many observational studies have reported numerous environment factors involved in myopia development, including education level, near work, urbanization, season of birth, and outdoor activities ^18-21^. Excessive near work including study, reading, and screen use were associated with having or developing myopia ^22^. Of these, the influence of electronic products use time remains controversial. While some studies reported a significant association between increased computer and smart phone use time and myopia development ^23, 24^, a meta-analysis reported s no association of screen time with the prevalence or incidence of myopia ^25^. Recent studies have shown that children who increased time spent outdoors (but not indoors) have a lower possibility of incident myopia ^26, 27^. Despite intensive efforts and numerous studies, there remains no consensus regarding primary myopia risk factors and effective prevention methods, in part due to the lack of human intervention study for the difficulty in implementing behavioral modification at large-scale samples. Thus, longitudinal studies of myopia development, particularly human behavioral intervention study on large scale are urgently needed.

We initiated the Myopic Epidemiology and Intervention Study (MEIS) in January 2019with support from the State Key Laboratory of Ophthalmology, Optometry and Visual Science, National Clinical Research Center for Ocular Disease under leadership of National Health Commission of China. The MEIS was designed to examine refraction-confirmed myopia among millions of schoolchildren in Wenzhou City with semiannual examination for ten years, taking into consideration of gender, educational level, educational system, and region of habitation. Until now, we have completed three test points of myopia: June 2019, December 2019, and June 2020, which provide a large-scale longitudinal study of 12-month follow-up, including the precious data for 6-month before and after the coronavirus disease (COVID-19) epidemic.

Since the outbreak of COVID-19 was first reported in late December 2019 in Wuhan, Hubei province, China ^28^, it has quickly caused an unprecedented global pandemic ^27^. Governments from many countries have imposed various strict containment measures on citizens including case isolation, limited outdoor activities, and school closures to prevent COVID-19 virus spread ^29^. According to the data from UNESCO, more than 87% of the world’s student population from > 160 countries are affected by these lockdown measures ^30^. COVID-19 quarantine has led to substantial changes in daily life among school-going children, including sharply dropped schooling times, but the increased online digital screen times and decreased outdoor activity times^31^. This provides unique and unprecedented opportunity to identify and evaluate the effect of behavioral modifications reeling from COVID-19 quarantine on the incidence and/or the worsening of myopia among schoolchildren ^32^.

Wenzhou is a coastal city located in Zhejiang province in China with 9.25 million local residents, of which many are usually businessmen by profession and frequently work and live in different cities including Wuhan. There were approximately 180,000 Wenzhou people working in Wuhan and 33,000 of them returned from Wuhan after COVID-19 outbreak, which led to Wenzhou was the worst affected city outside Hubei province in China in early February 2020. Therefore, schoolchildren in Wenzhou were strictly isolated to study at home from January to May 2020. It provided a practical experimental model to identify the critical risk factors for behavioral modification of developing myopia.

To the best of our knowledge, no comprehensive evaluation of the influence of COVID-19 quarantine on myopia development was reported. In the current longitudinal study across over one-million schoolchildren, we aimed to determine the prevalence of myopia and assess the impact of COVID-19 quarantine on myopia progression and incidence. The one-million mega dataset of myopia general survey allows to investigate the prevalence and to identify risk factors of myopia at the grade (even at birth month) scale. Specifically, we sought to address two critical questions: (1) Was six-month behavioral modifications during COVID-19 quarantine sufficient to change myopia progression course? (2) What are key risk factors for the occurrence and progression of myopia especially high myopia? - The findings from our study not only provides the first human behavioral intervention data at the one million-scale to identify the grade-specific causal factors, but also offer valuable information for the development of evidence-based myopia prevention and control strategies for children by school grade level.

## Methods

### Study design and participants

As part of our ongoing efforts within the MEIS project, the current survey of refraction-confirmed myopia was conducted among more than one-million schoolchildren in Wenzhou City, Zhejiang Province, China in June 2019, and followed by two examinations in December 2019 and June 2020. To facilitate this comprehensive myopia survey, we developed a semi-automated vision examination, and an information-entry pipeline, the Adolescent Myopia Information Management System (AMIMS), which involves manual inspection of automated refractometry data, automatic data import, and collaboration between clinicians and a statistician (Supplemental Figure S1).

In June of 2019, a census of 1,305 elementary and high schools encompassing different educational systems (key school and non-key school [i.e., public, sport, and martial arts]) in 11 districts of Wenzhou was conducted. There were 1,060,925 students aged 7 to 18 years recruited at the baseline, including 612,648 elementary school students (grades 1-6 in Chinese education system), 264,661 junior high school students (grades 7-9), and 183,616 senior high school students (grades 10-12). All students identified in the census were invited to complete a self-administered questionnaire regarding demographic information and their knowledge of myopia. Certified technicians were trained at the Wenzhou Medical University Affiliated Eye Hospital with respect to standard procedures for determining visual acuity (VA) and auto refraction testing. Each school in the district was equipped with an auto refractometer (GoldEye RM-9000) and electronic logarithmic visual chart (GoldEye CM-1900C) by the Wenzhou Municipal Government for assessing the degree of myopia. These resources were utilized by trained technicians to examine all participants. Among the 1,060,925 students, 18,627 students did not attend the VA and auto refraction testing. Participants with refraction values deemed nonstandard by the optometry staff were also excluded (N = 26,350), as were participants with atypical personal identity information (N = 14,119), resulting in 1,001,749 eligible participants (94.4% of the initial study population) remaining in the study (Figure 1).

**Figure 1.**
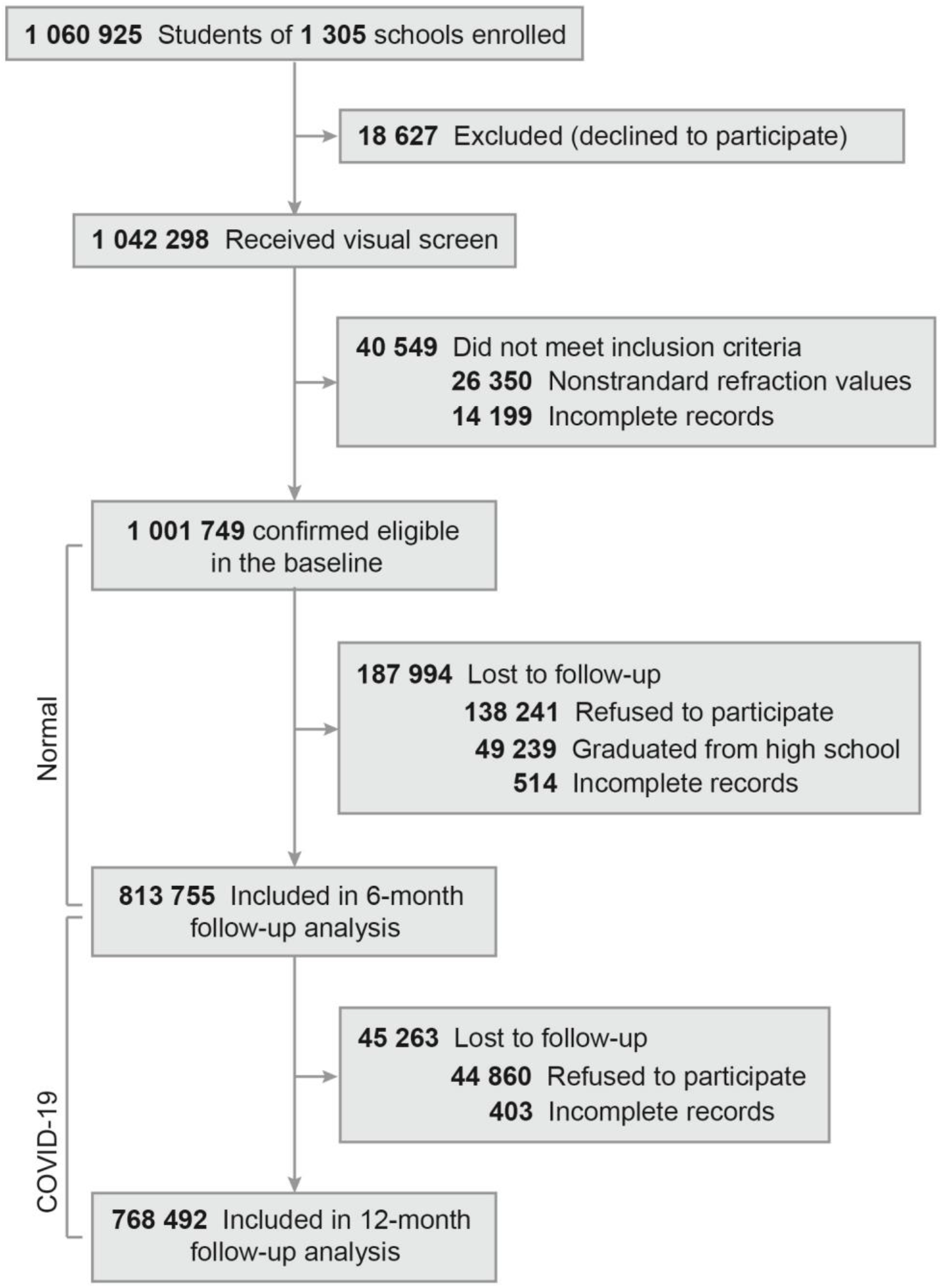
Flow of participants in the current longitudinal study with 12-month follow-up.

Subsequently, two follow-up examinations were conducted in December 2019 involving 813,755 students (81.2%) and June 2020 involving 768,492 students (76.7%). It was not possible to follow the remaining participants mainly because they had graduated from high school or refused to participate.

The present study was approved by the Ethics Committee of the Wenzhou Medical University Affiliated Eye Hospital (approval numbers Wmu191204 and Wmu191205). All procedures were conducted in accordance with institutional/national research committee ethical standards as well as the 1964 Helsinki declaration. Participant completion of the self-administered questionnaire was considered informed consent.

### Measurements and Outcomes

Due to the large sample size, performing the criterion standard of cycloplegic refraction to diagnose myopia was not applicable. Thus, the current survey adopted an alternative method of VA and autorefraction testing to identify myopia. VA was evaluated using an “E-type” standard logarithmic visual chart at a distance of 5 m. All students underwent noncycloplegic refraction testing using an automated refractometer, followed by complement subjective refraction testing for validation using an “E-type” standard logarithmic visual chart. Students who required corrective lenses were examined with and without eyeglasses, and naked eye refraction data were used to calculate the spherical equivalent. Myopia was identified as an uncorrected VA of less than 20/25, and spherical equivalent refraction (SER) was equaled to (sphere + [cylinder/2]) of -0.5 diopters (D) or less^33^. Since analysis for right and left eyes yielded a satisfactory correlation (Pearson correlation coefficient = 0.954, Supplemental Figure S2), SER data from the right eye were arbitrarily used for assessing myopia development, as reported in previous studies ^34, 35^. An SER of -6.0 D or less indicated as high myopia, and an SER between -0.5 D and -6.0 D defined as non-high myopia ^36^.

From June 2019 to December 2019, the first 6-month interval indicated as “Normal” period without the influence of COVID-19 quarantine, and the second 6-month interval from January 2020 to June 2020 indicated as “COVID-19” period with the influence of COVID-19 quarantine. Since COVID-19 quarantine had a great influence on alterations of schoolchildren lifestyle and study behaviors, it provided a practical intervention model for comparing the differences of risk factors related to myopia between normal period and COVID-19 period. Earlier studies ^37-39^ have used the 6-month interval to measure myopia progression for exploring the intervention strategies. In addition, during August 2020, we further used a new-designed questionnaire to survey 12,013 students, which were randomly selected from all grades. The questionnaire consisted of 17 questions, including the basic characteristics of schoolchildren, myopia information of their parents and siblings, and students’ lifestyle or learning approaches such as outdoor activities times and online course times during normal period and COVID-19 period.

### Statistical Analysis

The prevalence of myopia and high myopia were calculated by all eligible participants in three examinations. For prevalence, 95% confidence intervals (CIs) were calculated according to the procedure reported by Newcombe ^40^. To examine factors associated with the incidence of myopia, the Cox proportional hazard regression analysis was performed on students who attended all three examinations and did not have myopia at the baseline. Similarly, Cox proportional hazard regression analysis was conducted to reveal risk factors associated with the incidence of high myopia on students who attended all three examinations and did not have high myopia at the baseline. These multivariate Cox regression analyses were adjusted for grade, age, gender, birth month, educational system (key/non-key school), and habitation (urban/rural). The *survival* package in R was applied to perform multivariate Cox regression analyses.

The significance of differences between categorical variables was assessed using the *Fisher’s* test, and that of differences between continuous variables was assessed using Student’s *t* test. Two-sided P values were used in all statistical analyses. The Pearson correlation analysis was used to calculate the correlation of SER between right- and left-eye. The R software (ver. 3.6.1) was used for all statistical analyses (http://www.r-project.org).

## Results

### The prevalence of myopia and high myopia

We enrolled and collected data from a total of 1,060,925 students at the baseline (June 2019). After stringent sample filtering, 1,001,749 students (550,756 males and 450,993 females) were included in the current study (Figure 1 and Supplemental Table S1). There were 580,177 students in elementary schools, 250,839 in junior high schools, and 170,733 in senior high schools, covering almost all schools (1,305/1,336 = 97.8%) in 11 districts of Wenzhou City (Supplemental Figure S3). Of the 1,001,749 students, 813,755 (81.2%) were reexamined after the first six months (Normal period) and 768,492 (76.7%) after the second six months (COVID-19 period) (Figure 1). At the baseline, the myopia prevalence was 52.89% (95% CI, 52.79% to 52.99%), and the high myopia prevalence was 4.11% (95% CI, 4.07% to 4.15%). Table 1 shows the prevalence of myopia and high myopia among schoolchildren across three examinations categorized by gender, educational level, educational system, and habitation. The high prevalence of myopia among schoolchildren is similar with other cities of China and East Asia countries (Supplemental Table S2).

**Table 1.** Prevalence of myopia and its severity among Chinese schoolchildren.

| Characteristic<br>s | Prevalence (95% CI) |  |  |  |  |  |
| --- | --- | --- | --- | --- | --- | --- |
|  | Baseline<br>(Jun-2019) |  | 6-month follow-up<br>(Dec-2019) |  | 12-month follow-up<br>(Jun-2020) |  |
|  | Myopia | High myopia | Myopia | High myopia | Myopia | High myopia |
| **Total** | 52.89%<br>(52.79%-52.99%) | 4.11%<br>(4.07%-4.15%) | 53.9%<br>(53.79%-54.01%) | 4.24%<br>(4.2%-4.29%) | 59.35%<br>(59.24%-59.46%) | 4.99%<br>(4.94%-5.04%) |
| **Gender** |  |  |  |  |  |  |
| Male | 50.37%<br>(50.24%-50.51%) | 4.01%<br>(3.96%-4.07%) | 51.51%<br>(51.37%-51.66%) | 4.18%<br>(4.12%-4.24%) | 56.6%<br>(56.45%-56.75%) | 4.92%<br>(4.86%-4.99%) |
| Female | 55.96%<br>(55.82%-56.11%) | 4.23%<br>(4.17%-4.29%) | 56.79%<br>(56.63%-56.95%) | 4.32%<br>(4.25%-4.39%) | 62.67%<br>(62.51%-62.83%) | 5.07%<br>(4.99%-5.14%) |
| **Educational Level** |  |  |  |  |  |  |
| Elementary school | 35.41%<br>(35.29%-35.53%) | 0.82%<br>(0.8%-0.85%) | 34.38%<br>(34.24%-34.53%) | 0.72%<br>(0.69%-0.74%) | 42.82%<br>(42.67%-42.97%) | 1.12%<br>(1.08%-1.15%) |
| Junior high school | 74.01%<br>(73.84%-74.18%) | 6.32%<br>(6.23%-6.42%) | 71.71%<br>(71.53%-71.9%) | 5.48%<br>(5.38%-5.57%) | 75.25%<br>(75.06%-75.43%) | 6.78%<br>(6.67%-6.88%) |
| Senior high school | 81.26%<br>(81.07%-81.44%) | 12.03%<br>(11.87%-12.18%) | 81.68%<br>(81.49%-81.88%) | 12.21%<br>(12.05%-12.38%) | 82.57%<br>(82.38%-82.77%) | 13.34%<br>(13.17%-13.52%) |
| **Educational System** |  |  |  |  |  |  |
| Non-Key school | 51.39%<br>(51.28%-51.5%) | 3.49%<br>(3.45%-3.53%) | 52.16%<br>(52.04%-52.28%) | 3.51%<br>(3.47%-3.56%) | 57.98%<br>(57.86%-58.1%) | 4.37%<br>(4.32%-4.42%) |
| Key school | 58.69%<br>(58.48%-58.9%) | 6.52%<br>(6.41%-6.63%) | 60.88%<br>(60.64%-61.11%) | 7.18%<br>(7.05%-7.31%) | 64.92%<br>(64.68%-65.16%) | 7.5%<br>(7.37%-7.63%) |
| **Region of Habitation** |  |  |  |  |  |  |
| Rural | 47.36%<br>(47.11%-47.6%) | 2.85%<br>(2.76%-2.93%) | 48.35%<br>(48.08%-48.63%) | 2.99%<br>(2.9%-3.08%) | 53.36%<br>(53.08%-53.64%) | 3.66%<br>(3.56%-3.77%) |
| Urban | 55.66%<br>(55.55%-55.77%) | 4.62%<br>(4.57%-4.67%) | 56.38%<br>(56.26%-56.5%) | 4.72%<br>(4.66%-4.77%) | 62.05%<br>(61.92%-62.18%) | 5.49%<br>(5.43%-5.54%) |

### The important role of education burden in myopia development

At the baseline, cross-sectionally, the prevalence of myopia increased with grade level increasing (Table 1), and was 35.41% (95% CI, 35.29% to 35.53%) among elementary school (grades 1-6), 74.01% (95% CI, 73.84% to 74.18%) among junior high school (grades 7-9), and 81.26% (95% CI, 81.07% to 81.44%) among senior high school (grades 10-12). The high myopia prevalence was significantly higher among senior high school (12.03%; 95% CI, 11.87% to 12.18%) than that among elementary school (0.82%; 95% CI, 0.8% to 0.85%). A grade-stratified analysis indicated that myopia development could be categorized into two grade groups: Grade Group I (grades 1-6) representing myopia sensitive stage and Grade Group II (grades 7-12) representing high myopia sensitive stage (Supplemental Figure S4-S6). A high increase in myopia prevalence of 9.9% per advance in grade level was observed in Grade Group I, whereas the rate increase in myopia prevalence slowed to 2.4% per advance in grade level in Grade Group II (Supplemental Tables S3-S5). The prevalence of high myopia increased from 4.46% (95% CI: 4.32% to 4.6%) to 13.25% (95% CI: 12.95% to 13.55%) during advancement through Grade Group II (Supplemental Table S3). Interestingly, the prevalence of myopia increased in a stepwise manner by grade, as determined by sequential analyses stratified by the students’ birth month (Figure 2A, Supplemental Figure S7A-B, and Supplemental Table S6). Notably, an average increase in myopia prevalence of 8.54% (95% CI, 6.77% to 9.93%; P < 0.001) in the Grade Group I and 4.32% (95% CI, 0.69% to 7.42%; P < 0.001) in the Grade Group II was observed in students of the same age born in the August of the year compared with those born in the September (Supplemental Figures S8-S9 and Supplemental Table S6), consistent with policies regarding the age of school enrollment in China that students born in August went to school a year earlier. It provides a direct clue that grade increasing of education burden could play more effects on myopia occurrence rather than age.

**Figure 2.**
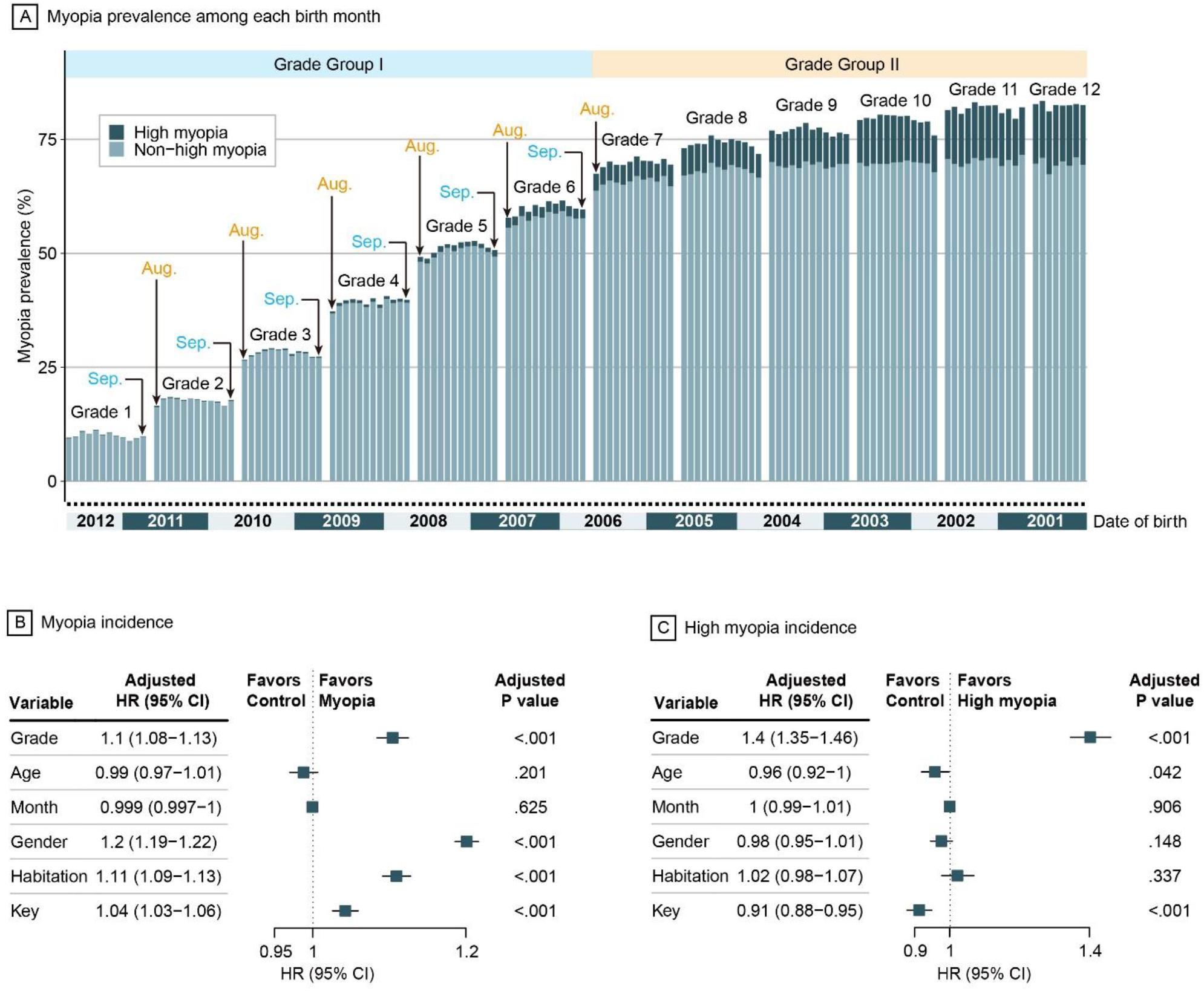
The prevalence and risk factors evaluation of myopia and high myopia. A) The prevalence of myopia and high myopia divided by schoolchildren birth month among all grades at the baseline. Each grade was divided into 12 bars according to birth month, ranging from September to August of next year according to Chinese enrollment policy. B) Forest plot shows risk factors for myopia based on multivariate Cox model. C) Forest plot shows risk factors for high myopia based on multivariate Cox model. In both Cox models, adjusted predictors included grade, age, gender, birth month, educational system, and habitation.

Consistently, by performing multivariate Cox regression analyses with adjustment, we found grade was statistically significantly associated with myopia (HR: 1.1, 95% CI, 1.08 to 1.13, P < 0.001) and high myopia (HR: 1.4, 95% CI, 1.35 to 1.46, P < 0.001), whereas age and birth month showed non-significant association with myopia and high myopia (Figure 2B-C). Furthermore, we found gender, habitation, and educational system showed a statistically significant association with myopia (Figure 2B). Compared with males, non-key school, and rural habitation separately, factors of females (HR: 1.2, 95% CI, 1.19 to 1.22, P < 0.001), key school (HR: 1.04, 95% CI, 1.03 to 1.06, P < 0.001), and urban habitation (HR: 1.11, 95% CI, 1.09 to 1.13, P < 0.001) had a higher risk to develop myopia.

### The influence of COVID-19 quarantine on myopia progression

During the COVID-19 pandemic, we found students’ online time (> 2h) substantially increased (Figure 3A) and outdoor activity time (> 1h) largely decreased (Figure 3B). For students in Grade Group I, the online time increased by 3.4 times from 22.7% in normal period to 77.5% in COVID-19 period, and the outdoor activity time decreased from 41.8% in normal period to 36.6% in COVID-19 period. With regard to students in Grade Group II, the online time increased by 2.07 times from 46.9% in normal period to 96.9% in COVID-19 period, and the outdoor activity time decreased by 1.71 times from 40.1% in normal period to 23.5% in COVID-19 period (Figure 3B). These increased online time and decreased outdoor time among school-aged children may lead to occur or worsen myopia.

**Figure 3.**
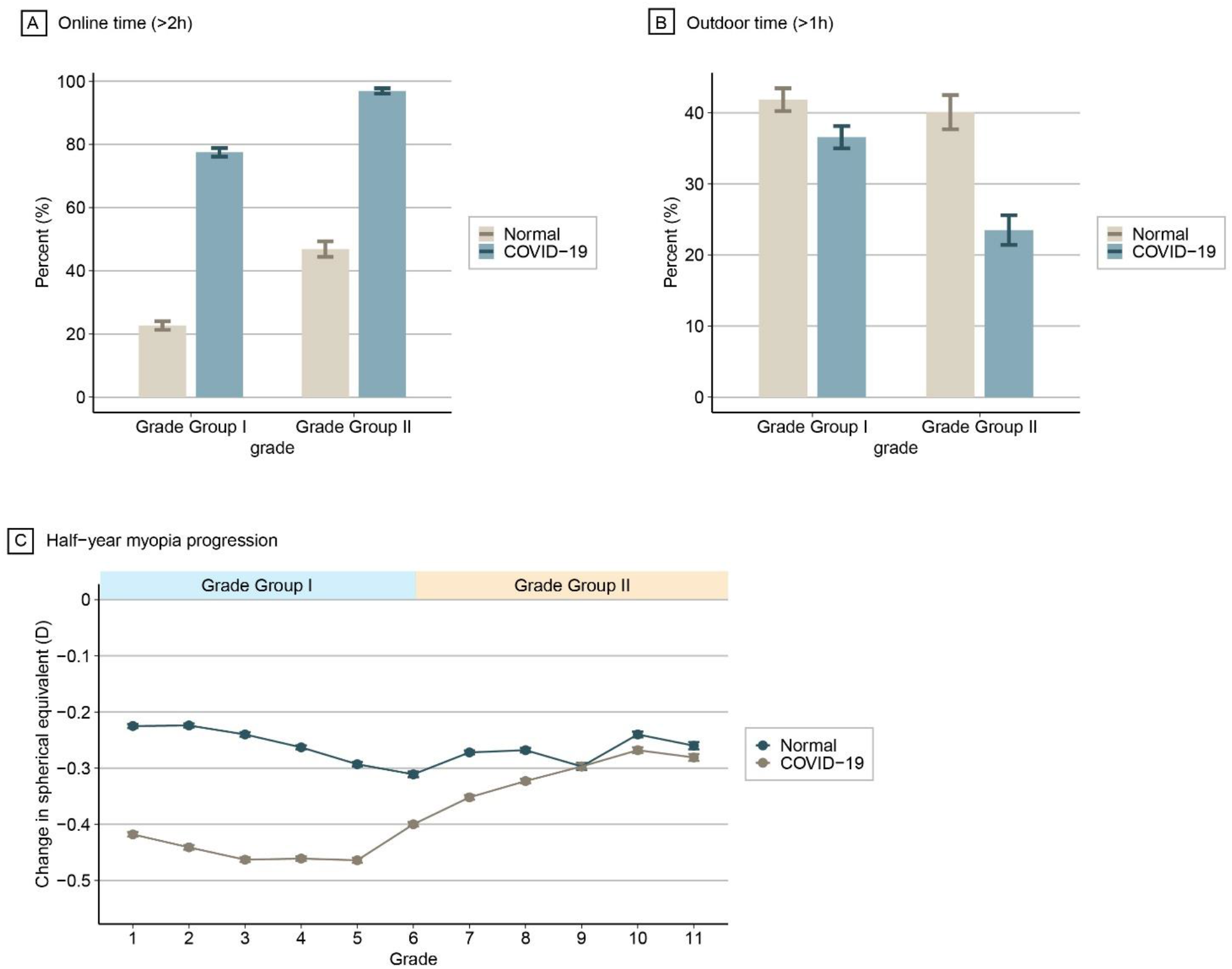
The behavioral modifications of schoolchildren during COVID-19 quarantine and its influence on myopia progression. A) The differences of schoolchildren’s online time between normal and COVID-19 period categorized by Grade Group I and II. B) The differences of schoolchildren’s outdoor activity time between normal and COVID-19 period categorized by Grade Group I and II. C) The differences of grade-specific half-year myopia progression between normal and COVID-19 period.

To assess the impact of COVID-19-induced lifestyle changes on myopia development, we found myopia progression speed among all schoolchildren increased of approximate 1.5 times from -0.263 D (95% CI, -0.262 to -0.264) during normal period to -0.39 D (95% CI, -0.389 to -0.391) during COVID-19 period (P < 0.001, Supplemental Table S7). Stratified by gender, educational level, educational system, and habitation, we also observed a significant exacerbation of myopia progression during COVID-19 period (P < 0.001, Supplemental Table S7). Notably, compared with normal period, we found that the faster myopia progression of students from Grade Group I than ones from Grade Group II during COVID-19 period (Figure 3C and Supplemental Table S8).

### The influence of COVID-19 quarantine on myopia incident

For all schoolchildren, we found that the half-year incidence rate of myopia increased from 8.5% in normal period to 9.87% in COVID-19 period (OR: 1.18, 95% CI, 1.15 to 1.20; Figure 4A). Stratified by female/male, key/non-key school, and urban habitation, schoolchildren had a significantly higher half-year incidence rate during COVID-19 period than that during normal period (P < 0.001, Figure 4A). COVID-19 quarantine showed no significant impact on half-year incidence rate among students from rural habitation (P = 0.62). With regard to educational level, students in Grade Group I showed a remarkably higher risk of developing myopia during COVID-19 period than normal period (OR: 1.36, 95% CI, 1.33 to 1.40; P < 0.001, Figure 4A), whereas students in Grade Group II had a prominently lower risk of incident myopia during COVID-19 period than normal period (OR: 0.45, 95% CI, 0.43 to 0.48; P < 0.001, Figure 4A). Interestingly, we found a distinct pattern that the half-year incidence rate of myopia during COVID-19 period consistently increased with grade increasing in Grade Group I, and the myopia half-year incidence rate during COVID-19 period gradually decreased with grade increasing in Grade Group II (Figure 4B and Supplemental Table S9). These results suggested COVID-19 was a risk factor for developing myopia among students in Grade Group I, whereas it was a protective factor for developing myopia among students in Grade Group II.

**Figure 4.**
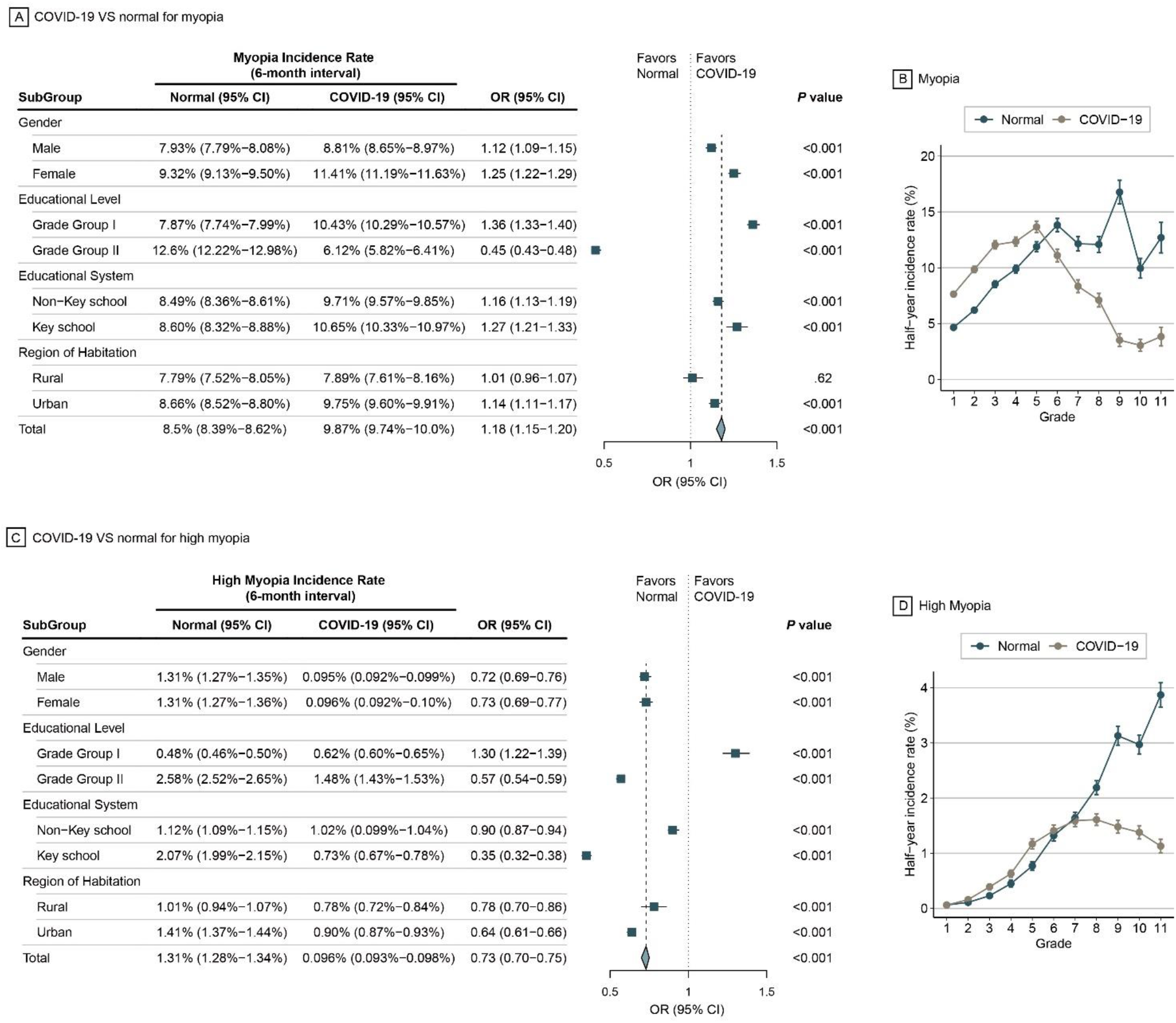
Significant changes in myopia incidence under the COVID-19 quarantine model. A) Forest plot indicates the differences of myopia incidence between normal and COVID-19 period stratified by gender, educational level, educational system, and habitation. B) The differences of grade-specific half-year myopia incidence between normal and COVID-19 period. C) Forest plot indicates the differences of high myopia incidence between normal and COVID-19 period stratified by gender, educational level, educational system, and habitation. B) The differences of grade-specific half-year high myopia incidence between normal and COVID-19 period.

As for high myopia, the half-year incidence rate among all schoolchildren decreased from 1.31% during normal period to 0.096% during COVID-19 period (OR: 0.73, 95% CI, 0.70 to 0.75; Figure 4C). Among students in Grade Group I, the half-year incidence rate of high myopia during COVID-19 period was significantly higher than that during normal period (OR: 1.30, 95% CI, 1.22 to 1.39, Figure 4C), whereas there was a lower half-year incidence rate of students in Grade Group II during COVID-19 period than that during normal period (OR: 0.57, 95% CI, 0.54 to 0.59, Figure 4C). Consistent with the identified COVID-19-influenced pattern of myopia, half-year incidence rate of high myopia during COVID-19 period gradually increased with grade increasing in Grade Group I, but decreased with grade increasing in Grade Group II (Figure 4D). In contrast, the half-year incidence rate of high myopia during normal period tended to persistently increase with grade increasing from 0.063% to 3.87% (Supplemental Table S10). Our finding consistently indicated COVID-19 contributed risk to incident high myopia among students in Grade Group I and had a protective effect on developing high myopia among students in Grade Group II.

## Discussion

### The COVID-19 quarantine reveals the crucial risk factors and uncovers the sufficiency of behavioral modifications and grade-specific control of myopia development

The ongoing COVID-19 pandemic has caused a public health emergency and global threat ^27^ and led to the population worldwide experiencing life-altering challenges. The COVID-19 quarantine also offers an unprecedented opportunity to evaluate the effect of behavioral modifications on the incidence and/or the worsening of myopia among schoolchildren on mega scale (such as one-million schoolchildren in the MEIS study). There existed positive and negative impacts of COVID-19 pandemic on various aspects of lifestyle behaviors ^31, 41, 42^. Hu et al. ^41^ reported approximately 90% of the Chinese participants had longer screen time and approximately 40% of the participants had inactive leisure-time physical exercise. Another study ^31^ on Chinese children and adolescents from five schools in Shanghai also reported that screen time remarkably increased by about 30 hours per week on average during the COVID-19 period. Consistently, we found that online time during COVID-19 period considerably increased by 3.4 times among Chinese students in Grade Group I and 2.07 times among ones in Grade Group II. Meanwhile the outdoor activity time substantially decreased among students among both Grade Groups I and II. These two factors (i.e. increased online time and decreased outdoor activity time) have been documented to enhance the risk of developing myopia ^23, 24, 27^. In light of COVID-19 quarantine led to billions of students are simultaneously exposed to the influence of cumulative risk factors for myopia ^32^, it is highly valuable to establish the COVID-19 quarantine intervention model in a real world study to uncover the crucial risk factors of myopia development.

First, some early studies ^37, 38^ may show that behavioral modifications for six months can influence myopia development. However, these studies are limited by sample size and by the difficulty in implementing behavioral modification at large scale. As the result, the sufficiency of behavioral modification pattern and time remain unclear. To the best of our knowledge, the EMIS study provides the first human intervention evidence at one-million scale to demonstrate that behavioral modification for six months during COVID-19 quarantine was sufficient to alter myopia progression. This sufficiency time for behavioral modification to be effective is essential in guiding the formation of myopia development and control among schoolchildren. Additional studies are needed to evaluate and demonstrate whether the COVID-19 pandemic exert lasting effects on the progression of myopia or high myopia (i.e. this specific pattern would persist during or after the COVID-19 quarantine) and to identify the shortest time for behavioral modification (such as three months) that is effective in influencing myopia development.

Furthermore, previous studies have demonstrated that myopia onset and peak progression are rapid in childhood, particularly around puberty ^43^. Compared with normal period, we found the COVID-19 quarantine had grade-specific effects on worsening the progression of myopia among schoolchildren, particularly among students in elementary school (Grade Group I), suggesting that elementary children were more vulnerable to be influenced by altered external environment resulting from COVID-19 quarantine. Interestingly, we uncovered a distinct pattern that COVID-19 quarantine was a risk factor of developing myopia or high myopia among students from Grade Group I, whereas COVID-19 quarantine was a protective factor for the development of myopia or high myopia among students from Grade Group II. Although COVID-19 increased the online time and decreased the outdoor activity time for all schoolchildren, COVID-19 still decreased the incidence rate of myopia and high myopia among students in Grade Group II, which may be due to the largely reduced study pressure.

Third, genetic factors play an important role in development of high myopia ^22, 44^. As result the extent of high myopia can be modified by environmental factors such as behavioral modification in COVID-19 quarantine is largely unknown. Interestingly, we found that behavioral modification increased the risk of developing high myopia in Grade I group and decreased the risk of incident high myopia in Grade II group. This study provide the clearest data yet in support of the environmental factors contribution to high myopia, which is collaborated by previous studies ^22, 45^.

### The EMIS study provide the first human behavioral intervention data at the one million-scale to identify the grade-specific causal factors for myopia prevention and control

Within our EMIS study population, the overall prevalence of myopia and high myopia were 52.89% and 4.11%, respectively. The prevalence of myopia among students of senior high school were 81.26%, which was similar to that reported among students of high school in 2010 in Shandong Province, China (84.6%) ^46^, Beijing, China (80.07%) ^2, 47^, Singapore (74.2%) ^48^, and South Korea (80.2%) ^49^. The prevalence of high myopia among all myopia cases at the end of senior high school was up to 13.25%, which is consistent with other Asian countries or regions. The prevalence of high myopia in Taiwan college freshmen ^50^ represented 26% of all myopia cases in 1988; by 2005, high myopia represented 40% of all myopia cases. Lin et al. ^51^ reported that 21% of students in Taiwan aged 18 years had high myopia in 2000, up from 10.9% in 1983. Notably, the prevalence of high myopia increased rapidly in Grade Group II of the EMIS study, indicating that although the onset of new cases of myopia slowed in high school, existing cases progressed in severity.

While several studies have evaluated and showed positive correlation between age and grade level with myopia development ^18, 52, 53^. However, these studies are limited by sample size and lack of the detailed analysis of these risk factors, rendering the importance of these risk factors uncertain.

In our survey of over one-million students, over 5,000 participants for each birth month, enabling stratification of the results by birth month. In students of the same age, differences in myopia were observed between those born in August and September, indicating that educational grade is more closely associated with development of myopia than age. Multivariate Cox regression analyses further validated the significant association between grade and myopia as well as high myopia after adjusting for other risk factors. Thus, our MEIS study with one-million participants establishes that educational burden could play key roles in myopia and high myopia.

Based on our multivariate Cox models, we found that female students had a higher risk of developing myopia or high myopia than male students, which was consistent with previous studies reporting a positive correlation between female sex and increased myopia risk in China ^47, 54, 55^. This difference could have a biological explanation related to hormonal differences, as postulated in a review of myopia in opposite-sex twins ^56^. In addition, traditional cultural factors might influence the behaviors of Chinese girls, such as Chinese girls prefer to white skin and avoid sun exposure, and are encouraged to be gentle and quiet, which may predispose girls to stay indoors more ^57^. We also observed that students from key school or urban habitation yielded a significantly higher risk of developing myopia or high myopia than those from non-key school or rural habitation. Consistently, Jan et al. ^55^ reported the prevalence of visual impairment was higher among Chinese students living in urban than ones living rural areas. More economically developed regions in China, such as Zhejiang, Jiangsu, and Shanghai, had a higher risk of visual impairment than less developed regions regardless of survey year ^55^.

The limitations of this study should be noted. First, the study participants were schoolchildren and did not include adults at universities and middle-aged or quinquagenarian individuals; hence our findings are limited to younger population. Furthermore, student’s online time and outdoor activity time changes during COVID-19 period were based on self-reported, thus these measurements might exist recall bias. Second, only Chinese schoolchildren were participants. In view of the ongoing COVID-19 spread worldwide, more studies are warranted to explore the influence of COVID-19 on myopia progression of schoolchildren based on more different ethnicities.

In summary, the present study is the largest longitudinal survey to assess the impact of COVID-19 quarantine-related lifestyle changes on myopia development among schoolchildren. We found an interesting COVID-19-induced pattern of developing myopia among children and adolescents, indicating different intervention strategies should be applied to control myopia among elementary and high schools. Our results should help inform educational and health policy decision making and the development of programs aimed at the prevention and control of myopia. Over the next decade, the MEIS will continue to support surveys of myopia in students twice annually, with a focus on discerning the effects of genetics, nutrition, and learning environment using questionnaire surveys and medical intervention studies to promote nationwide strategies for the prevention and control of myopia.

## Data Availability

The authors are open to sharing statistical codes and providing descriptive data in table form. Requests should be made to Jianzhong Su

## Contributors

J. Qu, J. Su and F. Lu conceived and designed the study; L. Xu, Y. Ma, Y. Zhang, H. Wang, G. Zhang, C. Tu, X. Liu, H. Chen, F. Chen, Y. Xiong, Z. Xue, X. Lu, M. Zhou and J. Sun performed myopia examination. Y. Ma, J. Yuan, G. Zhang, Y. Zhang, X. Lu, J. Li, W. Li, N. Wu, J. Su and L. Xu performed the statistical analyses and system development; Y. Ma, J. Chen, J. Su, L. Xu, J Qu, and F. Lu drafted the manuscript. All authors contributed to data collection, analysis, and interpretation.

## Acknowledgments

This work was supported by the Key Program of National Natural Science Foundation of China (81830027) to J. Qu; the National Natural Science Foundation of China (61871294), Zhejiang Provincial Natural Science Foundation of China (LR19C060001), and the Scientific Research Foundation for Talents of Wenzhou Medical University (QTJ18023) to J. Su; the Key Research and Development Program of Zhejiang Province (2020C03036) to L. Xu

## References

1. Holden BA, Fricke TR, Wilson DA, et al. Global prevalence of myopia and high myopia and temporal trends from 2000 through 2050. Ophthalmology 2016; 123(5): 1036–42.

2. You QS, Wu LJ, Duan JL, et al. Prevalence of myopia in school children in greater Beijing: the Beijing Childhood Eye Study. Acta ophthalmologica 2014; 92(5): e398–e406.

3. Ye S, Liu S, Li W, Wang Q, Xi W, Zhang X. Associations between anthropometric indicators and both refraction and ocular biometrics in a cross-sectional study of Chinese schoolchildren. BMJ open 2019; 9(5): e027212.

4. He M, Huang W, Zheng Y, Huang L, Ellwein LB. Refractive error and visual impairment in school children in rural southern China. Ophthalmology 2007; 114(2): 374-82. e1.

5. Ma Y, Qu X, Zhu X, et al. Age-specific prevalence of visual impairment and refractive error in children aged 3–10 years in Shanghai, China. Investigative ophthalmology & visual science 2016; 57(14): 6188–96.

6. Pi L-H, Chen L, Liu Q, et al. Refractive status and prevalence of refractive errors in suburban school-age children. International journal of medical sciences 2010; 7(6): 342.

7. Saw SM, Carkeet A, Chia KS, Stone RA, Tan DT. Component dependent risk factors for ocular parameters in Singapore Chinese children. Ophthalmology 2002; 109(11): 2065–71.

8. Yotsukura E, Torii H, Inokuchi M, et al. Current Prevalence of Myopia and Association of Myopia With Environmental Factors Among Schoolchildren in Japan. JAMA ophthalmology 2019; 137(11): 1233–9.

9. Jung SK, Lee JH, Kakizaki H, Jee D. Prevalence of myopia and its association with body stature and educational level in 19-year-old male conscripts in seoul, South Korea. Investigative ophthalmology & visual science 2012; 53(9): 5579–83.

10. Vitale S, Sperduto RD, Ferris FL, 3rd. Increased prevalence of myopia in the United States between 1971-1972 and 1999-2004. Archives of ophthalmology (Chicago, Ill : 1960) 2009; 127(12): 1632–9.

11. Bar Dayan Y, Levin A, Morad Y, et al. The changing prevalence of myopia in young adults: a 13-year series of population-based prevalence surveys. Investigative ophthalmology & visual science 2005; 46(8): 2760–5.

12. Knossow M, Gaudier M, Douglas A, et al. Mechanism of neutralization of influenza virus infectivity by antibodies. Virology 2002; 302(2): 294–8.

13. Gwiazda J, Hyman L, Dong LM, et al. Factors associated with high myopia after 7 years of follow-up in the Correction of Myopia Evaluation Trial (COMET) Cohort. Ophthalmic epidemiology 2007; 14(4): 230–7.

14. Resnikoff S, Pascolini D, Mariotti SP, Pokharel GP. Global magnitude of visual impairment caused by uncorrected refractive errors in 2004. Bulletin of the World Health Organization 2008; 86: 63–70.

15. Resnikoff S, Jonas JB, Friedman D, et al. Myopia–a 21st century public health issue. Investigative ophthalmology & visual science 2019; 60(3): Mi–Mii.

16. Knight JC, Udalova I, Hill AV, et al. A polymorphism that affects OCT-1 binding to the TNF promoter region is associated with severe malaria. Nat Genet 1999; 22(2): 145–50.

17. Goldschmidt E, Jacobsen N. Genetic and environmental effects on myopia development and progression. Eye 2014; 28(2): 126.

18. Ding B-Y, Shih Y-F, Lin LL, Hsiao CK, Wang I-J. Myopia among schoolchildren in East Asia and Singapore. survey of ophthalmology 2017; 62(5): 677–97.

19. He M, Xiang F, Zeng Y, et al. Effect of time spent outdoors at school on the development of myopia among children in China: a randomized clinical trial. Jama 2015; 314(11): 1142–8.

20. Rose KA, Morgan IG, Ip J, et al. Outdoor activity reduces the prevalence of myopia in children. Ophthalmology 2008; 115(8): 1279–85.

21. Pan CW, Ramamurthy D, Saw SM. Worldwide prevalence and risk factors for myopia. Ophthalmic & physiological optics : the journal of the British College of Ophthalmic Opticians (Optometrists) 2012; 32(1): 3–16.

22. Morgan IG, Ohno-Matsui K, Saw S-M. Myopia. The Lancet 2012; 379(9827): 1739–48.

23. Enthoven CA, Tideman JWL, Polling JR, Yang-Huang J, Raat H, Klaver CCW. The impact of computer use on myopia development in childhood: The Generation R study. Preventive medicine 2020; 132: 105988.

24. McCrann S, Loughman J, Butler JS, Paudel N, Flitcroft DI. Smartphone use as a possible risk factor for myopia. Clinical & experimental optometry 2020.

25. Lanca C, Saw SM. The association between digital screen time and myopia: A systematic review. Ophthalmic & physiological optics : the journal of the British College of Ophthalmic Opticians (Optometrists) 2020; 40(2): 216–29.

26. Jones LA, Sinnott LT, Mutti DO, Mitchell GL, Moeschberger ML, Zadnik K. Parental history of myopia, sports and outdoor activities, and future myopia. Investigative ophthalmology & visual science 2007; 48(8): 3524–32.

27. Sohrabi C, Alsafi Z, O’Neill N, et al. World Health Organization declares global emergency: A review of the 2019 novel coronavirus (COVID-19). International journal of surgery (London, England) 2020; 76: 71–6.

28. Zhu N, Zhang D, Wang W, et al. A Novel Coronavirus from Patients with Pneumonia in China, 2019. N Engl J Med 2020; 382(8): 727–33.

29. Parmet WE, Sinha MS. Covid-19 - The Law and Limits of Quarantine. N Engl J Med 2020; 382(15): e28.

30. Pellegrini M, Bernabei F, Scorcia V, Giannaccare G. May home confinement during the COVID-19 outbreak worsen the global burden of myopia? Graefe’s archive for clinical and experimental ophthalmology = Albrecht von Graefes Archiv fur klinische und experimentelle Ophthalmologie 2020; 258(9): 2069–70.

31. Xiang M, Zhang Z, Kuwahara K. Impact of COVID-19 pandemic on children and adolescents’ lifestyle behavior larger than expected. Progress in cardiovascular diseases 2020; 63(4): 531–2.

32. Wong CW, Tsai A, Jonas JB, et al. Digital Screen Time During COVID-19 Pandemic: Risk for a Further Myopia Boom? American journal of ophthalmology 2020.

33. Flitcroft DI, He M, Jonas JB, et al. IMI–Defining and classifying myopia: a proposed set of standards for clinical and epidemiologic studies. Investigative ophthalmology & visual science 2019; 60(3): M20–M30.

34. He M, Kong X, Chen Q, et al. Two-year changes in refractive error and related biometric factors in an adult Chinese population. JAMA ophthalmology 2014; 132(8): 978–84.

35. He M, Xiang F, Zeng Y, et al. Effect of Time Spent Outdoors at School on the Development of Myopia Among Children in China: A Randomized Clinical Trial. JAMA 2015; 314(11): 1142–8.

36. Knutsen LJ, Weiss SM. KW-6002 (Kyowa Hakko Kogyo). Curr Opin Investig Drugs 2001; 2(5): 668–73.

37. Jin JX, Hua WJ, Jiang X, et al. Effect of outdoor activity on myopia onset and progression in school-aged children in northeast China: the Sujiatun Eye Care Study. BMC ophthalmology 2015; 15: 73.

38. Huang PC, Hsiao YC, Tsai CY, et al. Protective behaviours of near work and time outdoors in myopia prevalence and progression in myopic children: a 2-year prospective population study. The British journal of ophthalmology 2020; 104(7): 956–61.

39. Nakamura T, Isogai N, Kojima T, Yoshida Y, Sugiyama Y. Posterior Chamber Phakic Intraocular Lens Implantation for the Correction of Myopia and Myopic Astigmatism: A Retrospective 10-Year Follow-up Study. American journal of ophthalmology 2019; 206: 1–10.

40. Newcombe RG. Two‐sided confidence intervals for the single proportion: comparison of seven methods. Statistics in medicine 1998; 17(8): 857–72.

41. Hu Z, Lin X, Chiwanda Kaminga A, Xu H. Impact of the COVID-19 Epidemic on Lifestyle Behaviors and Their Association With Subjective Well-Being Among the General Population in Mainland China: Cross-Sectional Study. Journal of medical Internet research 2020; 22(8): e21176.

42. Moore SA, Faulkner G, Rhodes RE, et al. Impact of the COVID-19 virus outbreak on movement and play behaviours of Canadian children and youth: a national survey. The international journal of behavioral nutrition and physical activity 2020; 17(1): 85.

43. Yip VC-H, Pan C-W, Lin X-Y, et al. The relationship between growth spurts and myopia in Singapore children. Investigative ophthalmology & visual science 2012; 53(13): 7961–6.

44. Guggenheim JA, Kirov G, Hodson SA. The heritability of high myopia: a reanalysis of Goldschmidt’s data. Journal of medical genetics 2000; 37(3): 227–31.

45. Wang SK, Guo Y, Liao C, et al. Incidence of and Factors Associated With Myopia and High Myopia in Chinese Children, Based on Refraction Without Cycloplegia. JAMA ophthalmology 2018; 136(9): 1017–24.

46. Wu JF, Bi HS, Wang SM, et al. Refractive error, visual acuity and causes of vision loss in children in Shandong, China. The Shandong Children Eye Study. PLoS One 2013; 8(12): e82763.

47. Wu LJ, You QS, Duan JL, et al. Prevalence and associated factors of myopia in high-school students in Beijing. PLoS One 2015; 10(3): e0120764.

48. Quek TP, Chua CG, Chong CS, et al. Prevalence of refractive errors in teenage high school students in Singapore. Ophthalmic and Physiological Optics 2004; 24(1): 47–55.

49. Rim TH, Kim S-H, Lim KH, Choi M, Kim HY, Baek S-H. Refractive errors in koreans: the Korea national health and nutrition examination survey 2008-2012. Korean Journal of Ophthalmology 2016; 30(3): 214–24.

50. Wang T, Chiang T-H, Wang T-H, Lin LL, Shih Y-F. Changes of the ocular refraction among freshmen in National Taiwan University between 1988 and 2005. Eye 2009; 23(5): 1168–9.

51. Lin LL-K, Shih Y-F, Hsiao CK, Chen C. Prevalence of myopia in Taiwanese schoolchildren: 1983 to 2000. Annals Academy of Medicine Singapore 2004; 33(1): 27–33.

52. Morgan IG, French AN, Ashby RS, et al. The epidemics of myopia: Aetiology and prevention. Progress in retinal and eye research 2018; 62: 134–49.

53. Williams KM, Bertelsen G, Cumberland P, et al. Increasing Prevalence of Myopia in Europe and the Impact of Education. Ophthalmology 2015; 122(7): 1489–97.

54. Guo K, Yang DY, Wang Y, et al. Prevalence of myopia in schoolchildren in Ejina: the Gobi Desert children eye study. Investigative ophthalmology & visual science 2015; 56(3): 1769–74.

55. Jan C, Xu R, Luo D, et al. Association of Visual Impairment With Economic Development Among Chinese Schoolchildren. JAMA pediatrics 2019; 173(7): e190914.

56. Miller EM. Reported myopia in opposite sex twins: a hormonal hypothesis. Optometry and vision science: official publication of the American Academy of Optometry 1995; 72(1): 34–6.

57. Lu B, Congdon N, Liu X, et al. Associations between near work, outdoor activity, and myopia among adolescent students in rural China: the Xichang Pediatric Refractive Error Study report no. 2. Archives of ophthalmology (Chicago, Ill : 1960) 2009; 127(6): 769–75.

