## Supplemental Tables and Figures for "COVID-19 Quarantine Reveals Grade-specific Behavioral Modification of Myopia: One-Million Chinese Schoolchildren Study"

### Supplemental Appendix

### Supplemental Figures

**Supplemental Figure S1.** The Adolescent Myopia Information Management System.

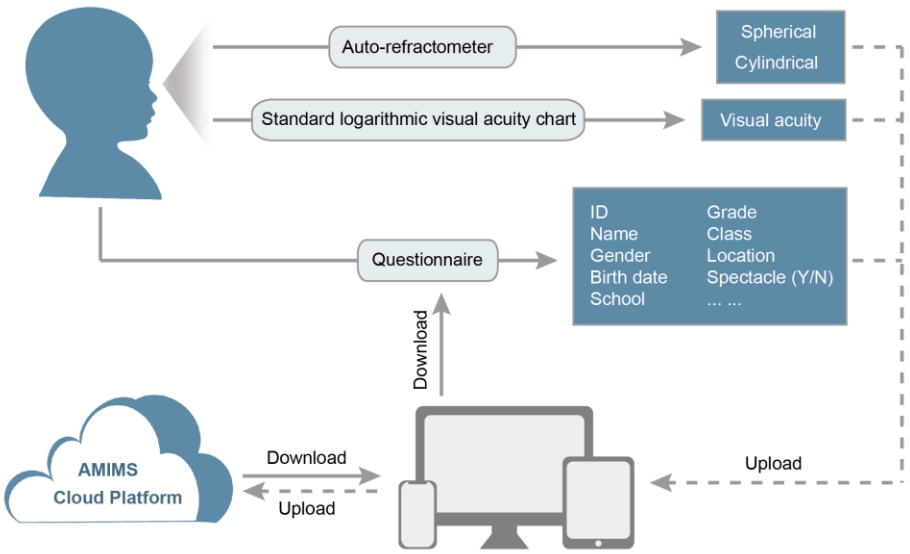

**Supplemental Figure S2. Correlations between left- and right-eyesight across three examinations.** A) the correlation at the baseline, B) the correlation at the 6-month follow-up, and C) the correlation at the 12-month follow-up. The average correlation coefficient (r) across three examinations is 0.954. OD (Oculus Dexter) represents right-eyesight and OS (Oculus Sinister) represents left-eyesight.

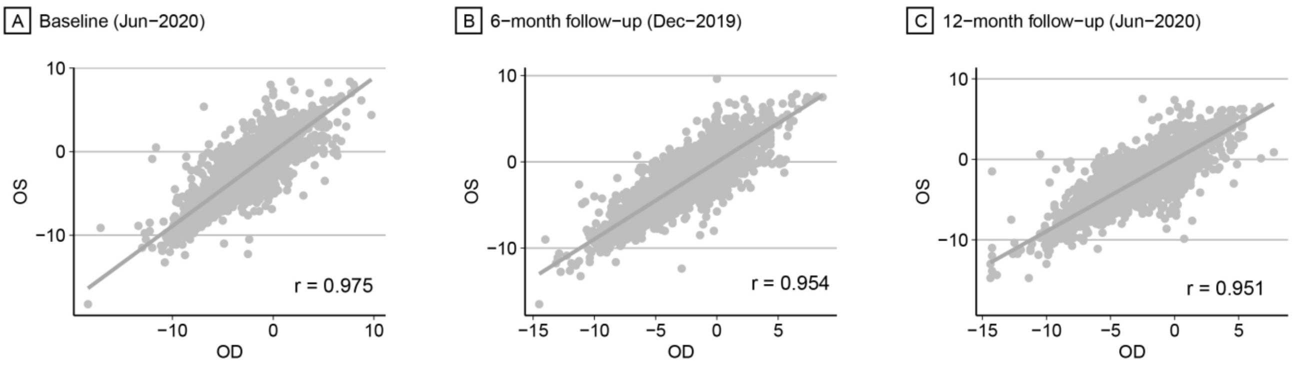

**Supplemental Figure S3. Population proportion of one-million children students in 11 main districts of Wenzhou City.** The color legend indicates the proportion of schoolchildren in each district among all participants.

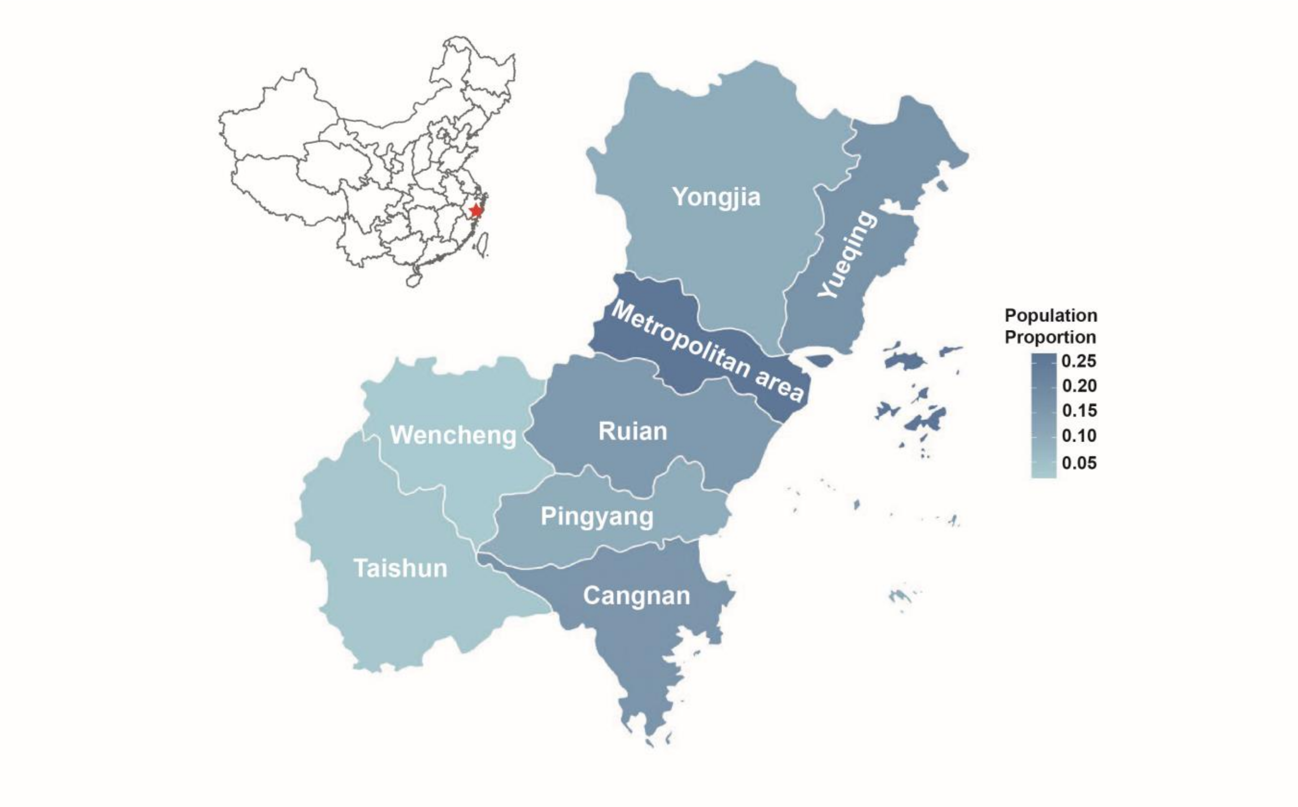

**Supplemental Figure S4. Grade-specific prevalence of myopia severity among Chinese schoolchildren at the baseline.** A) The grade-specific prevalence of overall, non-high, and high myopia among schoolchildren. B)-D) The grade-specific prevalence of overall, non-high, and high myopia amng schoolchildren sub-grouped by gender (male/female, B), habitation (Urban/rural, C) and educational system (key/non-key, D) Error bars indicate 95% confidence intervals.

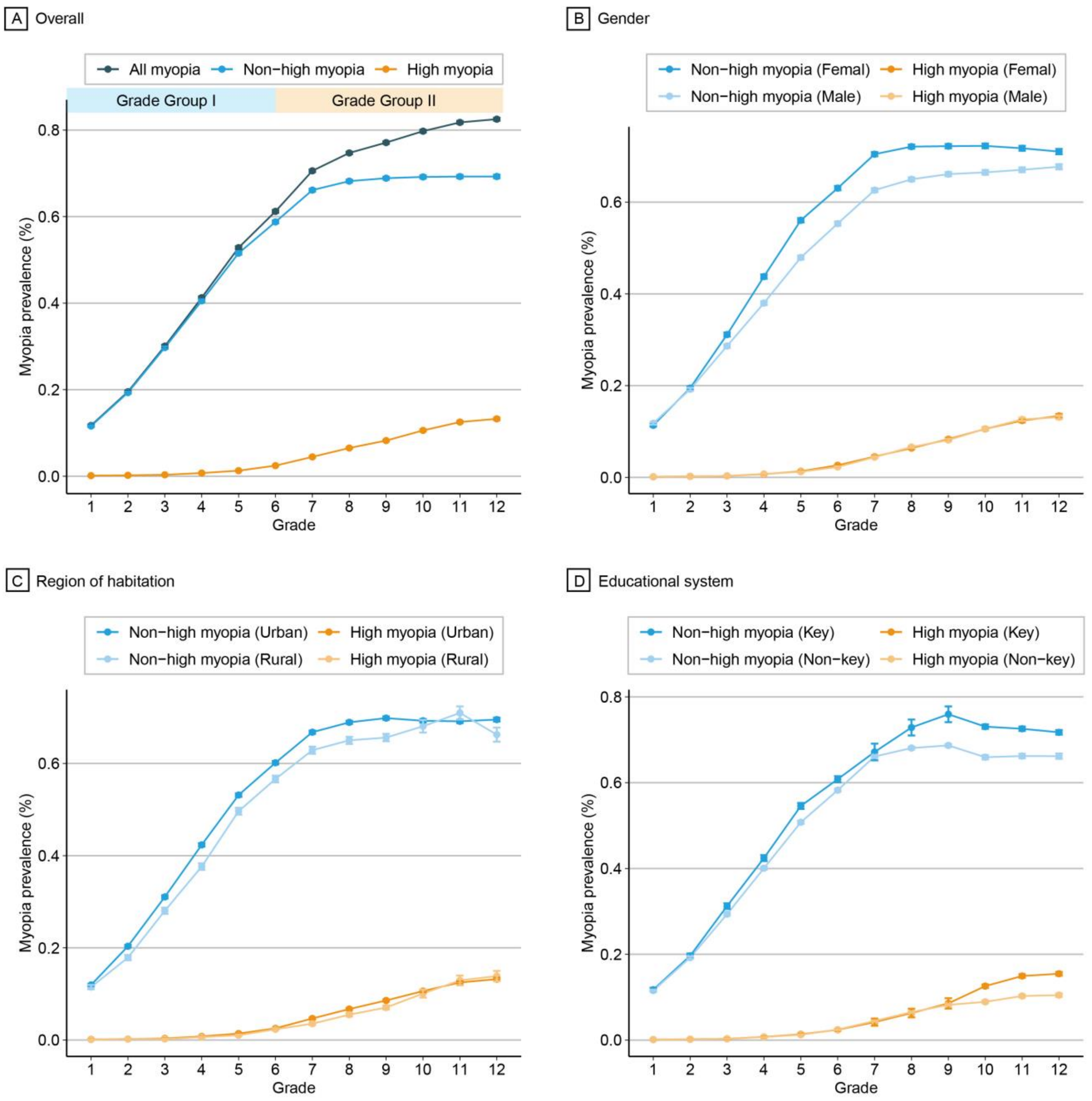

**Supplemental Figure S5. Grade-specific prevalence of myopia severity among Chinese schoolchildren at the 6-month follow-up.** A) The grade-specific prevalence of overall, non-high, and high myopia among schoolchildren. B)-D) The grade-specific prevalence of overall, non-high, and high myopia amng schoolchildren sub-grouped by gender (male/female, B), habitation (Urban/rural, C) and educational system (key/non-key, D). Error bars indicate 95% confidence intervals.

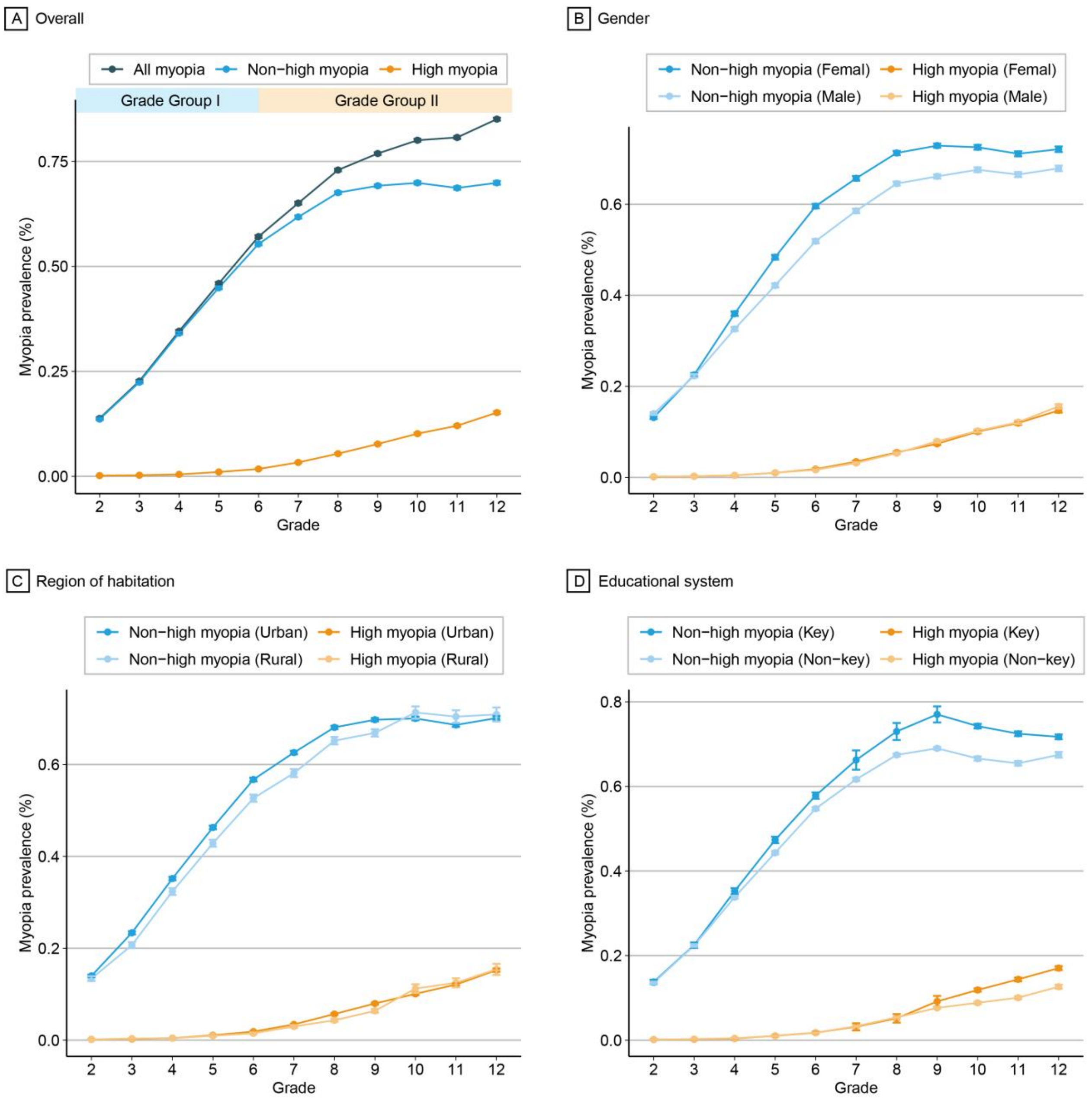

**Supplemental Figure S6. Grade-specific prevalence of myopia severity among Chinese schoolchildren at the 12-month follow-up.** A) The grade-specific prevalence of overall, non-high, and high myopia among schoolchildren. B)-D) The grade-specific prevalence of overall, non-high, and high myopia amng schoolchildren sub-grouped by gender (male/female, B), habitation (Urban/rural, C) and educational system (key/non-key, D). Error bars indicate 95% confidence intervals.

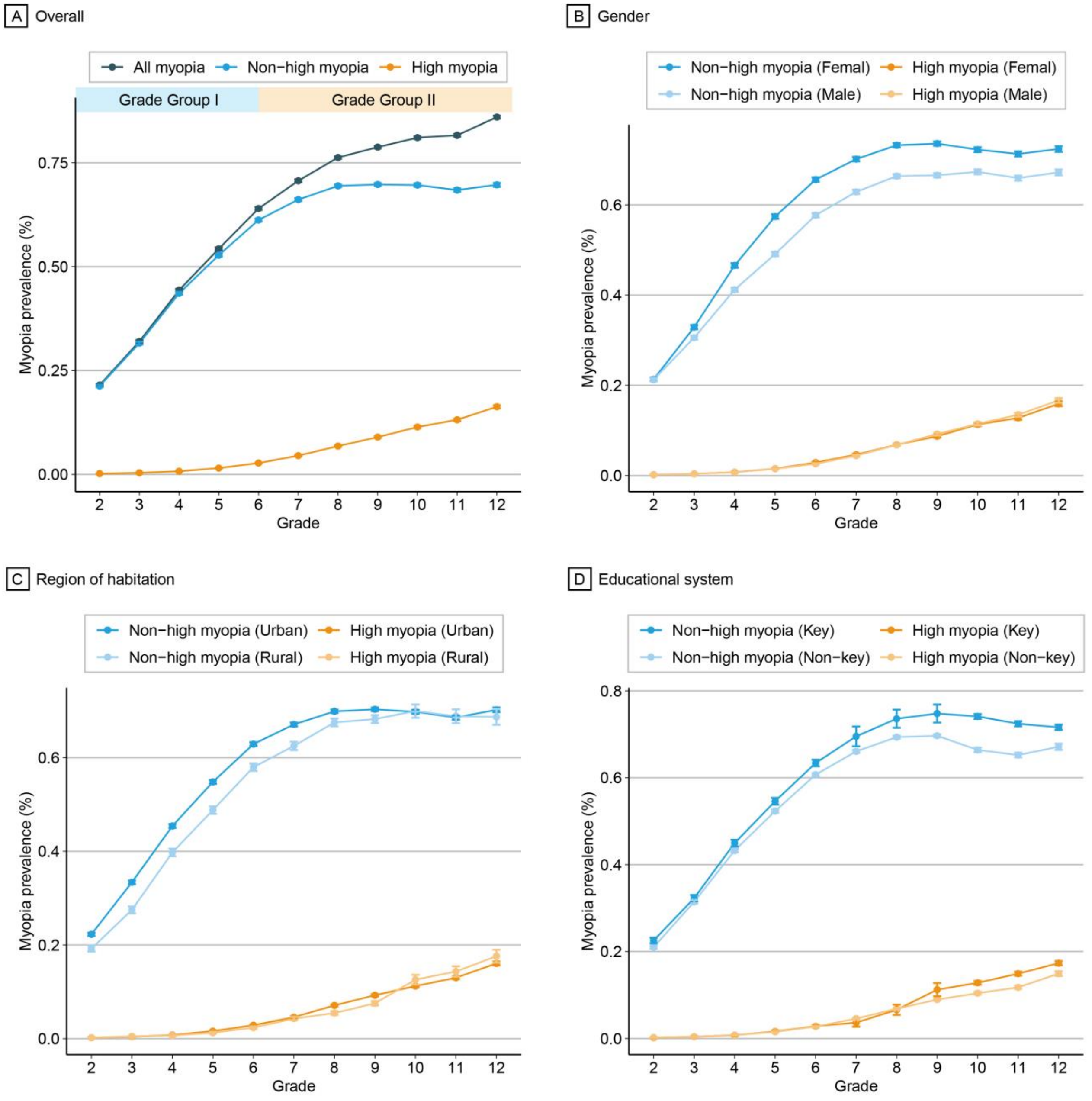

**Supplemental Figure S7. The prevalence of myopia and high myopia divided by schoolchildren birth month among all grades at the 6-month follow-up (A) and 12-month follow-up (B).** Each grade was divided into 12 bars according to schoolchildren birth months, ranging from September to August of next year according to Chinese enrollment policy.

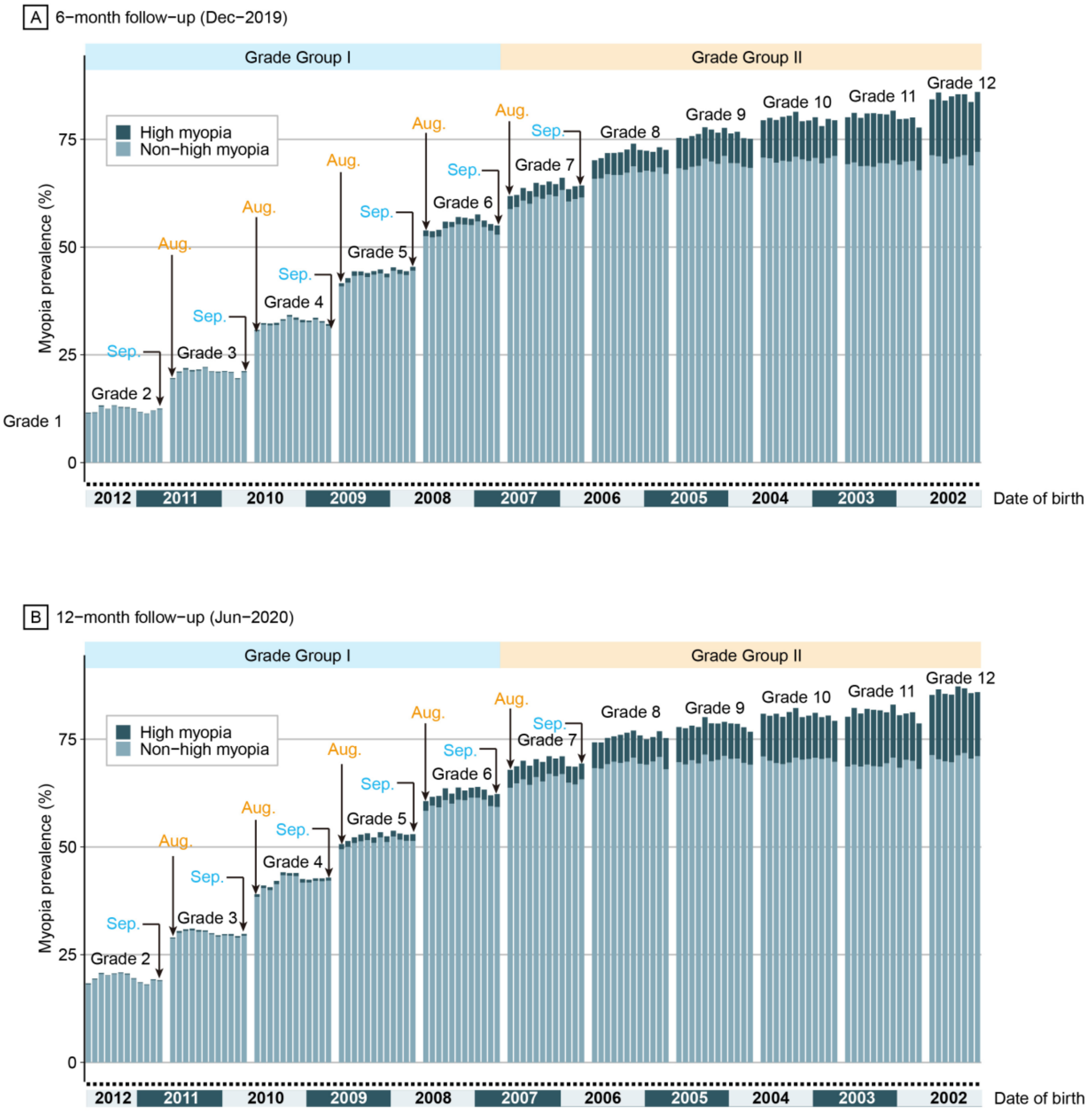

**Supplemental Figure S8. Differences in the prevalence of myopia between schoolchildren birth month of August and September of the same year among three examinations.** A) Baseline (June 2019), B) 6-month follow-up (December 2019), C) 12-month follow-up (June 2020).

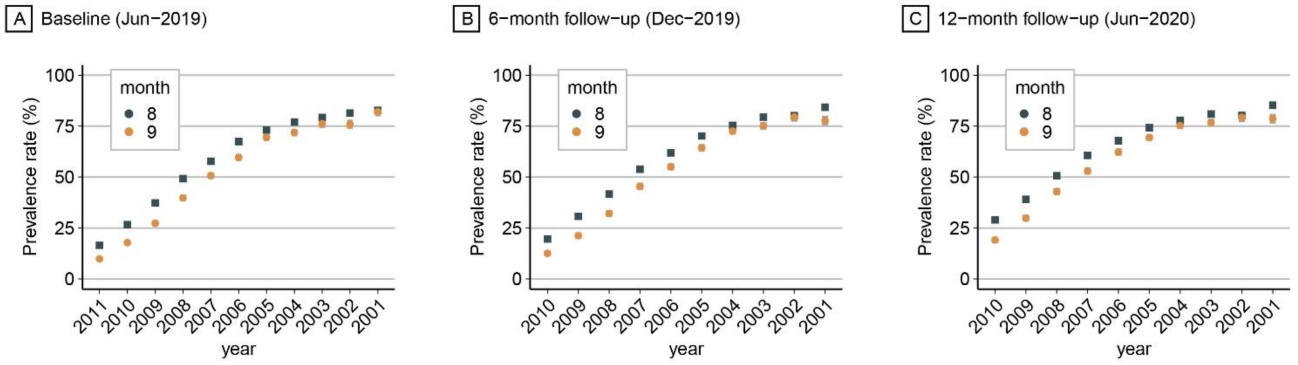

**Supplemental Figure S9. Differences in the prevalence of high myopia between schoolchildren birth month of August and September of the same year among three examinations.** A) Baseline (June 2019), B) 6-month follow-up (December 2019), C) 12-month follow-up (June 2020).

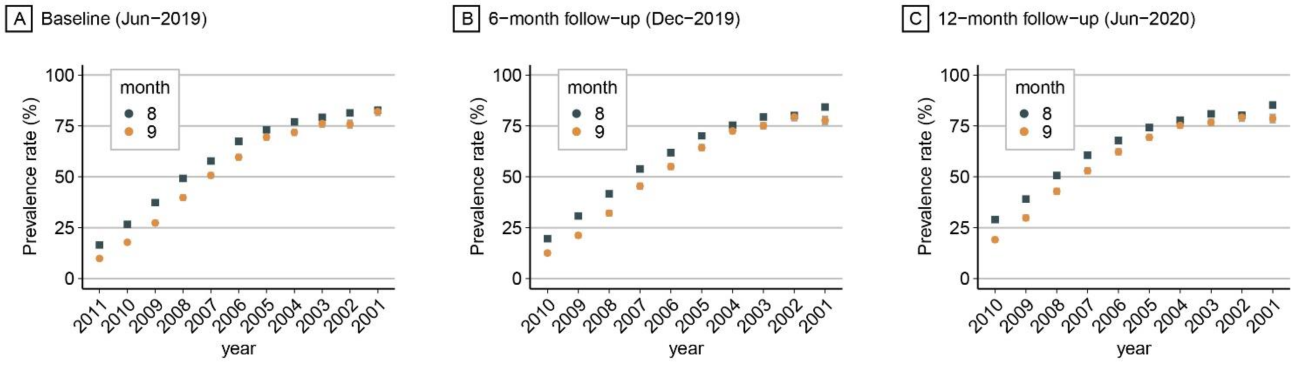

### Supplemental Tables

**Supplemental Table S1. Characteristics of school children across three time points used in the current survey**

| Characteristics | Baseline (Jun-2019) | | 6-month follow-up (Dec-2019) | | 12-month follow-up (Jun-2020) | |
|---|---|---|---|---|---|---|
| | Number of Samples | Proportion | Number of Samples | Proportion | Number of Samples | Proportion |
| **Total** | 1,007,149 | 100% | 813,755 | 100% | 768,492 | 100% |
| **Gender** | | | | | | |
| Male | 550,756 | 54.98% | 445,610 | 54.76% | 419,916 | 54.64% |
| Female | 450,993 | 45.02% | 368,145 | 45.24% | 348,576 | 45.36% |
| **Age (years)** | | | | | | |
| ⩽7 | 67,219 | 6.71% | 58,525 | 7.19% | NA | NA |
| 8 | 99,819 | 9.96% | 87,486 | 10.75% | 55,587 | 7.23% |
| 9 | 94,737 | 9.46% | 83,362 | 10.24% | 83,288 | 10.84% |
| 10 | 95,240 | 9.51% | 83,922 | 10.31% | 79,526 | 10.35% |
| 11 | 95,524 | 9.54% | 84,144 | 10.34% | 80,011 | 10.41% |
| 12 | 93,111 | 9.29% | 78,419 | 9.64% | 80,157 | 10.43% |
| 13 | 89,565 | 8.94% | 77,937 | 9.58% | 74,841 | 9.74% |
| 14 | 87,010 | 8.69% | 77,101 | 9.47% | 74,143 | 9.65% |
| 15 | 84,618 | 8.45% | 67,881 | 8.34% | 71,785 | 9.34% |
| 16 | 65,155 | 6.50% | 53,890 | 6.62% | 63,506 | 8.26% |
| 17 | 61,292 | 6.12% | 47,231 | 5.80% | 50,578 | 6.58% |
| ⩾18 | 68,459 | 6.83% | 13,859 | 1.70% | 55,073 | 7.17% |
| **Educational level** | | | | | | |
| Elementary school | 580,177 | 57.92% | 429,657 | 52.80% | 409,143 | 53.24% |
| Junior high school | 250,839 | 25.04% | 229,302 | 28.18% | 215,892 | 28.09% |
| Senior high school | 170,733 | 17.04% | 154,796 | 19.02% | 143,457 | 18.67% |
| **Educational system** | | | | | | |
| Non-Key school | 796,161 | 79.48% | 651,393 | 80.05% | 616,576 | 80.23% |
| Key school | 205,588 | 20.52% | 162,362 | 19.95% | 151,916 | 19.77% |
| **District of origin** | | | | | | |
| Yueqing | 170,014 | 16.97% | 130,215 | 16.00% | 125,472 | 16.33% |
| Pingyang | 96,852 | 9.67% | 83,793 | 10.30% | 77,991 | 10.15% |
| Wencheng | 24,201 | 2.42% | 20,675 | 2.54% | 19,501 | 2.54% |
| Yongjia | 98,138 | 9.80% | 78,615 | 9.66% | 74,860 | 9.74% |
| Taishun | 32,306 | 3.22% | 26,901 | 3.31% | 25,338 | 3.30% |
| Dongtou | 13,919 | 1.40% | 11,781 | 1.45% | 11,532 | 1.50% |
| Ruian | 150,404 | 15.01% | 123,515 | 15.18% | 117,482 | 15.29% |

| | | | | | | |
|---|---|---|---|---|---|---|
| Ouhai | 80,892 | 8.08% | 68,609 | 8.43% | 64,271 | 8.36% |
| Cangnan | 164,972 | 16.47% | 126,173 | 15.51% | 82,793 | 10.77% |
| Lucheng | 98,829 | 9.87% | 81,300 | 9.99% | 77,086 | 10.03% |
| Longwan | 71,222 | 7.11% | 62,178 | 7.64% | 92,166 | 12.00% |
| **Region of habitation** | | | | | | |
| Rural | 160,170 | 15.99% | 128,437 | 15.78% | 119,324 | 15.53% |
| Urban | 777,102 | 77.57% | 622,398 | 76.48% | 556,778 | 72.45% |
| Ambiguity | 64,477 | 6.44% | 62,920 | 7.73% | 92,390 | 12.02% |

**Supplemental Table S2. Prevalence of myopia among schoolchildren in East and Southeastern Asia**

| Locality | Year | Age | Participants | Prevalence (%) | Cut-off (D) | Reference |
|---|---|---|---|---|---|---|
| Beijing | 2014 | 7-18 | 15,066 | 9.7-74.2 | ≥ -0.5 | [1, 2] |
| Beijing | 2014 | 16-18 | 4,677 | 80.7 | ≥ -1.00 | [3] |
| ShunYi | 1998 | 5-15 | 5,884 | 45.9 | ≥ -0.5 | [4] |
| Chaoyang | 2011 | 5-14 | 4,249 | 7.4-67.5 | ≥ -0.5 | [5] |
| Tianjin | 2019 | 6-15 | 482 | 71.2 | ≥ -0.5 | [6] |
| Guangzhou | 2005 | 13-17 | 2,454 | 36.8-53.9 | ≥ -0.5 | [7] |
| Guangzhou | 2003 | 7-15 | 4,364 | 7.7-78.4 | ≥ -0.5 | [8] |
| Ejina | 2014 | 6-21 | 1,565 | 10-71.3 | ≥ -0.5 | [9] |
| Shandong | 2010 | 4-17 | 6,026 | 1.7-84.6 | ≥ -0.5 | [10] |
| Heilongjiang | 2009 | 5-18 | 1,675 | 0.9-8.2 | ≥ -0.5 | [11] |
| Yongchuan | 2007 | 6-15 | 3,070 | 0.42-27.11 | ≥ -0.5 | [12] |
| Neimenggu | 2003 | 7-17 | 1,057 | 5.8 | ≥ -0.5 | [13] |
| Xichang | 2007 | 11-17 | 1,892 | 62.3 | ≥ -0.5 | [14] |
| HongKong | 2000 | 6-15 | 7,560 | 17-53.1 | ≥ -0.5 | [15] |
| HongKong | 2010 | 6-12 | 2,651 | 18.3-61.5 | ≥ -0.5 | [16] |
| HongKong | 2001 | 13-15 | 1,191 | 62-87.2 | ≥ -0.5 | [17] |
| Taiwan | 2000 | 7-18 | 10,889 | 20-84 | ≥ -0.25 | [18] |
| Taiwan | 2000 | 7-18 | 45,345 | 5.8-84 | ≥ -0.25 | [19] |
| Taiwan | 2005 | 3-6 | 618 | 3.0-12.2 | ≥ -0.5 | [20] |
| Taiwan | 2005 | 12-19 | 20,609 | 70.3 | ≥ -0.5 | [21] |
| Korea | 2011 | 19 | 2,805 | 83.3 | ≥ -0.5 | [22] |
| Korea | 2012 | 5-19 | 7,486 | 20.4-80.2 | ≥ -0.75 | [23] |
| Korea | 2005 | 1-18 | 8,633 | 1.1-45.7 | ≥ -0.5 | [24] |
| Seoul | 2010 | 19 | 23,616 | 96.5 | ≥ -0.5 | [25] |
| Singapore | 2002 | 15-19 | 946 | 73.9 | ≥ -0.5 | [26] |
| Singapore | 2001 | 7-9 | 1,453 | 29-53.1 | ≥ -0.5 | [27] |
| Singapore | 2008 | 6-7 | 628 | 29.1 | ≥ -0.5 | [28] |

| | | | | | | |
|---|---|---|---|---|---|---|
| Japan | 1996 | 3-17 | 17,320 | 37.6-46.2 | ≥ -0.5 | [29] |
| Tokyo | 2017 | 6-14 | 1,416 | 63.1-94.2 | ≥ -0.5 | [30] |

**Supplemental Table S3. Prevalence of myopia and high myopia across every grade in the baseline (Jun-2019)**

| Grade group | Educational level | Grade | Count | Prevalence (95% CI) | |
|---|---|---|---|---|---|
| | | | | Myopia | High myopia |
| Grade Group I | Elementary school | 1 | 104669 | 11.73% (11.53%-11.92%) | 0.13% (0.11%-0.16%) |
| | | 2 | 98389 | 19.52% (19.28%-19.77%) | 0.2% (0.17%-0.23%) |
| | | 3 | 95543 | 30.06% (29.77%-30.35%) | 0.33% (0.29%-0.36%) |
| | | 4 | 96215 | 41.26% (40.95%-41.57%) | 0.73% (0.67%-0.78%) |
| | | 5 | 94442 | 52.82% (52.51%-53.14%) | 1.28% (1.21%-1.35%) |
| | | 6 | 90919 | 61.21% (60.89%-61.53%) | 2.44% (2.34%-2.54%) |
| Grade Group II | Junior high school | 7 | 87742 | 70.59% (70.29%-70.89%) | 4.46% (4.32%-4.6%) |
| | | 8 | 86103 | 74.72% (74.42%-75.01%) | 6.51% (6.35%-6.68%) |
| | | 9 | 76994 | 77.12% (76.82%-77.41%) | 8.23% (8.04%-8.42%) |
| | Senior high school | 10 | 61688 | 79.75% (79.43%-80.07%) | 10.58% (10.33%-10.82%) |
| | | 11 | 59806 | 81.78% (81.47%-82.09%) | 12.52% (12.25%-12.78%) |
| | | 12 | 49239 | 82.52% (82.19%-82.86%) | 13.25% (12.95%-13.55%) |

**Supplemental Table S4. Prevalence of myopia and high myopia across every grade in the 6-month follow-up (Dec-2019)**

| Grade group | Educational level | Grade | Count | Prevalence (95% CI) | |
|---|---|---|---|---|---|
| | | | | Myopia | High myopia |
| Grade Group I | Elementary school | 2 | 91049 | 13.79% (13.57%-14.02%) | 0.17% (0.14%-0.19%) |
| | | 3 | 86238 | 22.66% (22.38%-22.94%) | 0.26% (0.22%-0.29%) |
| | | 4 | 84212 | 34.55% (34.23%-34.87%) | 0.45% (0.41%-0.5%) |
| | | 5 | 84851 | 45.93% (45.6%-46.27%) | 1.02% (0.96%-1.09%) |
| | | 6 | 83307 | 57.09% (56.76%-57.43%) | 1.75% (1.66%-1.84%) |
| Grade Group II | Junior high school | 7 | 74088 | 65.07% (64.72%-65.41%) | 3.3% (3.17%-3.43%) |
| | | 8 | 79188 | 72.96% (72.65%-73.27%) | 5.39% (5.23%-5.55%) |
| | | 9 | 76026 | 76.89% (76.59%-77.19%) | 7.68% (7.49%-7.87%) |
| | Senior high school | 10 | 57341 | 80.05% (79.72%-80.38%) | 10.15% (9.9%-10.4%) |
| | | 11 | 54377 | 80.72% (80.39%-81.05%) | 12.04% (11.77%-12.32%) |
| | | 12 | 43078 | 85.08% (84.74%-85.41%) | 15.17% (14.83%-15.51%) |

**Supplemental Table S5. Prevalence of myopia and high myopia across every grade in the 12-month follow-up (Jun-2020)**

| Grade group | Educational level | Grade | Count | Prevalence (95% CI) | |
|---|---|---|---|---|---|
| | | | | Myopia | High myopia |
| Grade Group I | Elementary school | 2 | 86491 | 21.51% (21.24%-21.78%) | 0.21% (0.18%-0.24%) |
| | | 3 | 82136 | 32.01% (31.69%-32.33%) | 0.41% (0.36%-0.45%) |
| | | 4 | 80378 | 44.35% (44.01%-44.7%) | 0.78% (0.72%-0.84%) |
| | | 5 | 80766 | 54.31% (53.97%-54.66%) | 1.54% (1.45%-1.62%) |

| | | | | | |
|---|---|---|---|---|---|
| | | 6 | 79372 | 64% (63.66%-64.33%) | 2.75% (2.64%-2.87%) |
| **Grade Group II** | Junior high school | 7 | 70855 | 70.67% (70.34%-71.01%) | 4.54% (4.38%-4.69%) |
| | | 8 | 75118 | 76.28% (75.97%-76.58%) | 6.83% (6.65%-7.01%) |
| | | 9 | 69919 | 78.78% (78.47%-79.08%) | 8.99% (8.78%-9.21%) |
| | Senior high school | 10 | 54161 | 81.05% (80.72%-81.38%) | 11.42% (11.15%-11.69%) |
| | | 11 | 51130 | 81.62% (81.28%-81.95%) | 13.17% (12.88%-13.46%) |
| | | 12 | 38166 | 86.01% (85.66%-86.36%) | 16.31% (15.93%-16.68%) |

**Supplemental Table S6. Prevalence of myopia and high myopia among each month**

| | | | **Prevalence (95% CI)** | | | | | |
|---|---|---|---|---|---|---|---|---|
| | **Birth** | | **Baseline (Jun-2019)** | | **6-month follow-up (Dec-2019)** | | **12-month follow-up (Jun-2020)** | |
| **Grade** | **Year** | **Month** | **Myopia** | **High myopia** | **Myopia** | **High myopia** | **Myopia** | **High myopia** |
| 1 | 2012 | 8 | 9.53% (8.91%-10.15%) | 0.2% (0.1%-0.29%) | NA | NA | NA | NA |
| 1 | 2012 | 7 | 9.75% (9.11%-10.38%) | 0.12% (0.05%-0.19%) | NA | NA | NA | NA |
| 1 | 2012 | 6 | 11.01% (10.33%-11.69%) | 0.19% (0.1%-0.29%) | NA | NA | NA | NA |
| 1 | 2012 | 5 | 10.4% (9.73%-11.08%) | 0.09% (0.02%-0.15%) | NA | NA | NA | NA |
| 1 | 2012 | 4 | 11.25% (10.55%-11.96%) | 0.16% (0.07%-0.24%) | NA | NA | NA | NA |
| 1 | 2012 | 3 | 10.2% (9.56%-10.84%) | 0.15% (0.07%-0.23%) | NA | NA | NA | NA |
| 1 | 2012 | 2 | 10.65% (9.98%-11.31%) | 0.1% (0.03%-0.16%) | NA | NA | NA | NA |
| 1 | 2012 | 1 | 10.01% (9.4%-10.62%) | 0.1% (0.03%-0.16%) | NA | NA | NA | NA |
| 1 | 2011 | 12 | 9.57% (8.97%-10.17%) | 0.08% (0.02%-0.13%) | NA | NA | NA | NA |
| 1 | 2011 | 11 | 8.78% (8.2%-9.36%) | 0.11% (0.04%-0.17%) | NA | NA | NA | NA |
| 1 | 2011 | 10 | 9.37% (8.76%-9.97%) | 0.12% (0.05%-0.2%) | NA | NA | NA | NA |
| 1 | 2011 | 9 | 9.85% (9.19%-10.51%) | 0.13% (0.05%-0.21%) | NA | NA | NA | NA |
| 2 | 2011 | 8 | 16.5% (15.71%-17.3%) | 0.24% (0.13%-0.35%) | 11.58% (10.86%-12.31%) | 0.2% (0.1%-0.3%) | 18.34% (17.43%-19.24%) | 0.26% (0.14%-0.37%) |
| 2 | 2011 | 7 | 18.12% (17.27%-18.97%) | 0.21% (0.11%-0.32%) | 11.66% (10.93%-12.4%) | 0.15% (0.06%-0.24%) | 19.44% (18.51%-20.36%) | 0.21% (0.11%-0.32%) |
| 2 | 2011 | 6 | 18.45% (17.57%- | 0.2% (0.1%- | 13.25% (12.47%- | 0.26% (0.15%- | 20.76% (19.8%- | 0.26% (0.14%- |

| | | | | | | | | |
|---|---|---|---|---|---|---|---|---|
| | | | 19.33%) | 0.3%) | 14.04%) | 0.38%) | 21.72%) | 0.38%) |
| 2 | 2011 | 5 | 18.26% (17.38%-19.15%) | 0.22% (0.11%-0.33%) | 12.47% (11.69%-13.25%) | 0.1% (0.03%-0.18%) | 20.22% (19.25%-21.2%) | 0.12% (0.04%-0.21%) |
| 2 | 2011 | 4 | 17.82% (16.94%-18.69%) | 0.22% (0.11%-0.32%) | 13.28% (12.47%-14.09%) | 0.15% (0.06%-0.24%) | 20.64% (19.65%-21.63%) | 0.19% (0.08%-0.29%) |
| 2 | 2011 | 3 | 18.21% (17.37%-19.06%) | 0.11% (0.04%-0.19%) | 12.91% (12.15%-13.66%) | 0.18% (0.09%-0.28%) | 20.87% (19.93%-21.81%) | 0.18% (0.08%-0.28%) |
| 2 | 2011 | 2 | 17.92% (17.06%-18.78%) | 0.18% (0.09%-0.28%) | 12.85% (12.08%-13.62%) | 0.17% (0.07%-0.26%) | 20.62% (19.66%-21.57%) | 0.26% (0.14%-0.38%) |
| 2 | 2011 | 1 | 17.61% (16.79%-18.42%) | 0.18% (0.09%-0.27%) | 12.55% (11.83%-13.28%) | 0.14% (0.06%-0.22%) | 19.55% (18.67%-20.44%) | 0.16% (0.07%-0.25%) |
| 2 | 2010 | 12 | 17.69% (16.88%-18.5%) | 0.13% (0.05%-0.2%) | 11.75% (11.05%-12.45%) | 0.11% (0.04%-0.18%) | 18.59% (17.72%-19.46%) | 0.18% (0.09%-0.28%) |
| 2 | 2010 | 11 | 17.5% (16.71%-18.3%) | 0.22% (0.12%-0.31%) | 11.37% (10.67%-12.06%) | 0.09% (0.02%-0.15%) | 18.1% (17.24%-18.96%) | 0.16% (0.07%-0.24%) |
| 2 | 2010 | 10 | 16.47% (15.68%-17.26%) | 0.15% (0.07%-0.24%) | 12.08% (11.36%-12.8%) | 0.1% (0.03%-0.17%) | 19.28% (18.38%-20.17%) | 0.17% (0.08%-0.27%) |
| 2 | 2010 | 9 | 17.81% (16.95%-18.68%) | 0.27% (0.15%-0.38%) | 12.53% (11.74%-13.31%) | 0.18% (0.08%-0.27%) | 19.09% (18.13%-20.05%) | 0.2% (0.09%-0.31%) |
| 3 | 2010 | 8 | 26.64% (25.65%-27.64%) | 0.21% (0.11%-0.31%) | 19.58% (18.68%-20.49%) | 0.22% (0.11%-0.32%) | 28.99% (27.93%-30.06%) | 0.26% (0.14%-0.38%) |
| 3 | 2010 | 7 | 27.62% (26.59%-28.64%) | 0.26% (0.14%-0.37%) | 21.11% (20.15%-22.07%) | 0.26% (0.14%-0.38%) | 30.5% (29.39%-31.61%) | 0.42% (0.27%-0.58%) |
| 3 | 2010 | 6 | 28.26% (27.22%-29.31%) | 0.28% (0.16%-0.41%) | 21.92% (20.92%-22.93%) | 0.35% (0.21%-0.49%) | 30.9% (29.75%-32.04%) | 0.41% (0.26%-0.57%) |
| 3 | 2010 | 5 | 28.95% (27.88%-30.03%) | 0.38% (0.23%-0.53%) | 21.46% (20.45%-22.46%) | 0.33% (0.19%-0.47%) | 31.05% (29.9%-32.21%) | 0.41% (0.25%-0.57%) |
| 3 | 2010 | 4 | 29.16% (28.08%-30.24%) | 0.29% (0.16%-0.42%) | 21.58% (20.58%-22.58%) | 0.26% (0.14%-0.39%) | 30.73% (29.59%-31.88%) | 0.4% (0.25%-0.56%) |
| 3 | 2010 | 3 | 28.92% (27.89%-29.94%) | 0.21% (0.11%-0.32%) | 22.17% (21.2%-23.14%) | 0.16% (0.06%-0.25%) | 30.63% (29.53%-31.73%) | 0.28% (0.16%-0.41%) |
| 3 | 2010 | 2 | 29.15% (28.12%-30.18%) | 0.41% (0.27%-0.56%) | 21.16% (20.19%-22.12%) | 0.15% (0.06%-0.24%) | 29.98% (28.86%-31.09%) | 0.26% (0.14%-0.38%) |
| 3 | 2010 | 1 | 27.9% (26.93%-28.87%) | 0.4% (0.26%-0.54%) | 21.09% (20.16%-22.02%) | 0.2% (0.1%-0.3%) | 29.51% (28.44%-30.57%) | 0.33% (0.19%-0.46%) |
| 3 | 2009 | 12 | 28.49% (27.55%-29.44%) | 0.31% (0.19%-0.43%) | 21.23% (20.3%-22.15%) | 0.16% (0.07%-0.25%) | 29.78% (28.72%-30.84%) | 0.31% (0.18%-0.44%) |
| 3 | 2009 | 11 | 28.33% (27.37%-29.28%) | 0.32% (0.2%-0.43%) | 21.02% (20.12%-21.93%) | 0.21% (0.11%-0.31%) | 29.81% (28.77%-30.85%) | 0.43% (0.28%-0.58%) |
| 3 | 2009 | 10 | 27.26% (26.34%-28.19%) | 0.16% (0.08%-0.24%) | 19.57% (18.67%-20.47%) | 0.24% (0.13%-0.35%) | 29.31% (28.25%-30.37%) | 0.38% (0.24%-0.52%) |
| 3 | 2009 | 9 | 27.33% (26.31%-28.35%) | 0.3% (0.17%-0.42%) | 21.21% (20.22%-22.2%) | 0.24% (0.13%-0.36%) | 29.82% (28.69%-30.96%) | 0.41% (0.26%-0.57%) |

| | | | | | | | | |
|---|---|---|---|---|---|---|---|---|
| 4 | 2009 | 8 | 37.28% (36.18%-38.37%) | 0.5% (0.34%-0.66%) | 30.79% (29.68%-31.89%) | 0.3% (0.17%-0.43%) | 39.03% (37.84%-40.22%) | 0.62% (0.43%-0.81%) |
| 4 | 2009 | 7 | 39.12% (38%-40.24%) | 0.68% (0.49%-0.87%) | 32.35% (31.21%-33.48%) | 0.35% (0.21%-0.5%) | 41.06% (39.84%-42.29%) | 0.63% (0.43%-0.82%) |
| 4 | 2009 | 6 | 39.68% (38.51%-40.85%) | 0.71% (0.51%-0.91%) | 32.27% (31.11%-33.44%) | 0.43% (0.27%-0.6%) | 40.67% (39.42%-41.92%) | 0.69% (0.48%-0.9%) |
| 4 | 2009 | 5 | 39.94% (38.78%-41.11%) | 0.81% (0.6%-1.02%) | 32.43% (31.25%-33.61%) | 0.51% (0.33%-0.69%) | 42.13% (40.86%-43.39%) | 0.79% (0.56%-1.02%) |
| 4 | 2009 | 4 | 39.7% (38.53%-40.86%) | 0.66% (0.47%-0.86%) | 33.25% (32.07%-34.43%) | 0.33% (0.18%-0.47%) | 44.11% (42.83%-45.39%) | 0.67% (0.46%-0.88%) |
| 4 | 2009 | 3 | 38.76% (37.67%-39.85%) | 0.56% (0.4%-0.73%) | 34.26% (33.12%-35.4%) | 0.38% (0.23%-0.52%) | 43.94% (42.72%-45.16%) | 0.63% (0.44%-0.82%) |
| 4 | 2009 | 2 | 40.11% (39%-41.23%) | 0.73% (0.53%-0.92%) | 33.64% (32.51%-34.77%) | 0.51% (0.34%-0.68%) | 43.96% (42.75%-45.18%) | 0.75% (0.54%-0.96%) |
| 4 | 2009 | 1 | 38.74% (37.7%-39.78%) | 0.73% (0.55%-0.91%) | 33.09% (32.01%-34.17%) | 0.58% (0.4%-0.75%) | 42.55% (41.39%-43.71%) | 0.85% (0.63%-1.06%) |
| 4 | 2008 | 12 | 40.62% (39.61%-41.64%) | 0.64% (0.47%-0.8%) | 32.92% (31.86%-33.97%) | 0.34% (0.21%-0.47%) | 42.4% (41.27%-43.53%) | 0.64% (0.46%-0.82%) |
| 4 | 2008 | 11 | 39.81% (38.8%-40.82%) | 0.72% (0.54%-0.89%) | 33.6% (32.54%-34.67%) | 0.4% (0.26%-0.54%) | 42.71% (41.56%-43.86%) | 0.61% (0.43%-0.8%) |
| 4 | 2008 | 10 | 40.03% (38.99%-41.06%) | 0.67% (0.5%-0.84%) | 32.82% (31.78%-33.86%) | 0.33% (0.2%-0.46%) | 42.69% (41.57%-43.81%) | 0.65% (0.47%-0.84%) |
| 4 | 2008 | 9 | 39.77% (38.66%-40.88%) | 0.6% (0.43%-0.78%) | 32.14% (31.01%-33.28%) | 0.43% (0.27%-0.59%) | 42.88% (41.65%-44.11%) | 0.72% (0.51%-0.93%) |
| 5 | 2008 | 8 | 49.19% (48.06%-50.32%) | 1.07% (0.83%-1.3%) | 41.61% (40.42%-42.8%) | 0.74% (0.54%-0.95%) | 50.65% (49.41%-51.88%) | 1.18% (0.91%-1.45%) |
| 5 | 2008 | 7 | 48.81% (47.67%-49.96%) | 1.1% (0.87%-1.34%) | 42.78% (41.58%-43.98%) | 1.04% (0.79%-1.29%) | 51.37% (50.13%-52.61%) | 1.35% (1.07%-1.64%) |
| 5 | 2008 | 6 | 50.06% (48.89%-51.23%) | 0.97% (0.74%-1.2%) | 44.36% (43.1%-45.62%) | 1.05% (0.79%-1.31%) | 52.24% (50.95%-53.54%) | 1.34% (1.05%-1.64%) |
| 5 | 2008 | 5 | 51.58% (50.37%-52.8%) | 1.29% (1.01%-1.56%) | 44.34% (43.08%-45.6%) | 0.93% (0.69%-1.18%) | 52.86% (51.56%-54.15%) | 1.56% (1.23%-1.88%) |
| 5 | 2008 | 4 | 51.98% (50.75%-53.21%) | 0.84% (0.61%-1.07%) | 43.97% (42.71%-45.22%) | 0.93% (0.69%-1.17%) | 53.16% (51.88%-54.45%) | 1.59% (1.27%-1.92%) |
| 5 | 2008 | 3 | 51.79% (50.63%-52.95%) | 1.33% (1.07%-1.6%) | 44.42% (43.24%-45.61%) | 0.83% (0.61%-1.04%) | 52.22% (51%-53.43%) | 1.25% (0.98%-1.52%) |
| 5 | 2008 | 2 | 52.41% (51.27%-53.55%) | 1.29% (1.03%-1.54%) | 44.81% (43.61%-46.02%) | 0.94% (0.71%-1.18%) | 53.41% (52.18%-54.65%) | 1.23% (0.96%-1.5%) |
| 5 | 2008 | 1 | 52.53% (51.47%-53.58%) | 1.05% (0.84%-1.27%) | 43.85% (42.72%-44.97%) | 0.86% (0.65%-1.07%) | 52.46% (51.3%-53.62%) | 1.32% (1.05%-1.58%) |
| 5 | 2007 | 12 | 52.71% (51.66%-53.77%) | 1.12% (0.9%-1.35%) | 45.3% (44.2%-46.39%) | 0.82% (0.62%-1.01%) | 53.77% (52.64%-54.89%) | 1.37% (1.11%-1.63%) |
| 5 | 2007 | 11 | 52.08% (51.05%-53.11%) | 0.96% (0.76%-1.16%) | 44.73% (43.64%-45.82%) | 1% (0.78%-1.22%) | 53.12% (52%-54.24%) | 1.41% (1.15%-1.68%) |

| | | | | | | | | |
|---|---|---|---|---|---|---|---|---|
| 5 | 2007 | 10 | 51.22% (50.14%-52.31%) | 0.97% (0.76%-1.18%) | 44.39% (43.28%-45.5%) | 0.88% (0.67%-1.08%) | 52.83% (51.68%-53.98%) | 1.4% (1.13%-1.67%) |
| 5 | 2007 | 9 | 50.71% (49.55%-51.87%) | 1.42% (1.15%-1.7%) | 45.42% (44.21%-46.63%) | 0.92% (0.69%-1.15%) | 52.95% (51.71%-54.19%) | 1.57% (1.26%-1.88%) |
| 6 | 2007 | 8 | 57.78% (56.64%-58.92%) | 2.17% (1.83%-2.5%) | 53.86% (52.66%-55.05%) | 1.33% (1.05%-1.6%) | 60.63% (59.43%-61.83%) | 2.25% (1.89%-2.62%) |
| 6 | 2007 | 7 | 58.08% (56.93%-59.22%) | 1.96% (1.64%-2.28%) | 53.68% (52.47%-54.89%) | 1.41% (1.12%-1.69%) | 61.63% (60.42%-62.83%) | 2.03% (1.68%-2.38%) |
| 6 | 2007 | 6 | 60.38% (59.22%-61.54%) | 2.22% (1.87%-2.57%) | 54.01% (52.77%-55.25%) | 1.57% (1.26%-1.87%) | 61.84% (60.6%-63.08%) | 2.64% (2.23%-3.05%) |
| 6 | 2007 | 5 | 59.13% (57.94%-60.31%) | 1.97% (1.63%-2.3%) | 55.9% (54.62%-57.18%) | 1.54% (1.22%-1.86%) | 63.6% (62.32%-64.87%) | 2.74% (2.31%-3.18%) |
| 6 | 2007 | 4 | 60.61% (59.41%-61.82%) | 2.49% (2.1%-2.87%) | 55.84% (54.54%-57.14%) | 1.2% (0.91%-1.48%) | 62.37% (61.07%-63.67%) | 2.32% (1.92%-2.73%) |
| 6 | 2007 | 3 | 60.1% (58.97%-61.23%) | 2.29% (1.95%-2.64%) | 56.98% (55.76%-58.21%) | 1.71% (1.39%-2.03%) | 63.8% (62.59%-65.01%) | 2.84% (2.42%-3.26%) |
| 6 | 2007 | 2 | 61.4% (60.25%-62.54%) | 2.39% (2.03%-2.75%) | 56.8% (55.6%-58%) | 1.58% (1.28%-1.88%) | 63.1% (61.91%-64.3%) | 2.28% (1.91%-2.65%) |
| 6 | 2007 | 1 | 60.9% (59.82%-61.98%) | 2.15% (1.83%-2.47%) | 56.56% (55.45%-57.68%) | 1.45% (1.18%-1.72%) | 63.75% (62.64%-64.86%) | 2.31% (1.96%-2.65%) |
| 6 | 2006 | 12 | 61.56% (60.49%-62.62%) | 2.3% (1.97%-2.63%) | 57.58% (56.47%-58.69%) | 1.64% (1.35%-1.92%) | 63.94% (62.84%-65.04%) | 2.54% (2.18%-2.9%) |
| 6 | 2006 | 11 | 60.28% (59.2%-61.37%) | 2.2% (1.87%-2.52%) | 56.15% (55.07%-57.24%) | 1.51% (1.24%-1.78%) | 63.3% (62.22%-64.38%) | 2.37% (2.02%-2.71%) |
| 6 | 2006 | 10 | 59.84% (58.75%-60.94%) | 2.21% (1.89%-2.54%) | 55.35% (54.21%-56.5%) | 1.53% (1.24%-1.81%) | 61.98% (60.83%-63.13%) | 2.51% (2.14%-2.88%) |
| 6 | 2006 | 9 | 59.62% (58.41%-60.82%) | 1.92% (1.58%-2.25%) | 55.01% (53.79%-56.24%) | 2.13% (1.77%-2.48%) | 62.28% (61.06%-63.5%) | 2.99% (2.56%-3.42%) |
| 7 | 2006 | 8 | 67.47% (66.35%-68.59%) | 3.75% (3.3%-4.2%) | 61.84% (60.6%-63.07%) | 3.02% (2.58%-3.45%) | 67.89% (66.67%-69.11%) | 4.15% (3.63%-4.67%) |
| 7 | 2006 | 7 | 68.91% (67.82%-70.01%) | 3.81% (3.35%-4.26%) | 62.11% (60.87%-63.36%) | 2.84% (2.42%-3.27%) | 68.71% (67.5%-69.92%) | 3.96% (3.45%-4.47%) |
| 7 | 2006 | 6 | 70.11% (69%-71.22%) | 4.14% (3.66%-4.62%) | 63.73% (62.46%-64.99%) | 2.92% (2.48%-3.36%) | 70% (68.77%-71.23%) | 4.33% (3.78%-4.87%) |
| 7 | 2006 | 5 | 69.41% (68.28%-70.54%) | 3.91% (3.43%-4.38%) | 62.97% (61.68%-64.25%) | 2.92% (2.47%-3.36%) | 68.84% (67.57%-70.1%) | 4.42% (3.86%-4.98%) |
| 7 | 2006 | 4 | 69.39% (68.26%-70.52%) | 4.33% (3.83%-4.83%) | 64.91% (63.62%-66.2%) | 3.26% (2.78%-3.74%) | 70.38% (69.12%-71.64%) | 4.17% (3.62%-4.72%) |
| 7 | 2006 | 3 | 70.09% (69.04%-71.14%) | 4.33% (3.86%-4.8%) | 64.41% (63.19%-65.63%) | 3.21% (2.76%-3.66%) | 69.74% (68.54%-70.93%) | 4.56% (4.02%-5.1%) |
| 7 | 2006 | 2 | 71.27% (70.2%-72.35%) | 4.32% (3.84%-4.8%) | 65.2% (63.96%-66.45%) | 3.04% (2.59%-3.49%) | 71.06% (69.85%-72.27%) | 4.12% (3.58%-4.65%) |
| 7 | 2006 | 1 | 70.28% (69.24%-71.32%) | 4.09% (3.64%-4.54%) | 64.59% (63.43%-65.76%) | 2.82% (2.42%-3.22%) | 70.51% (69.38%-71.64%) | 4.09% (3.6%-4.58%) |

| | | | | | | | | |
|---|---|---|---|---|---|---|---|---|
| 7 | 2005 | 12 | 70.25% (69.22%-71.28%) | 3.69% (3.27%-4.11%) | 66.09% (64.95%-67.23%) | 2.85% (2.45%-3.25%) | 71.04% (69.93%-72.16%) | 4.11% (3.62%-4.6%) |
| 7 | 2005 | 11 | 69.65% (68.6%-70.7%) | 3.93% (3.49%-4.37%) | 63.45% (62.27%-64.62%) | 2.86% (2.45%-3.26%) | 68.72% (67.57%-69.88%) | 3.82% (3.34%-4.29%) |
| 7 | 2005 | 10 | 70.69% (69.62%-71.76%) | 3.74% (3.3%-4.19%) | 64.12% (62.95%-65.3%) | 3% (2.58%-3.42%) | 68.62% (67.46%-69.78%) | 4.11% (3.61%-4.6%) |
| 7 | 2005 | 9 | 69.47% (68.27%-70.66%) | 4.77% (4.21%-5.32%) | 64.31% (63.01%-65.61%) | 2.78% (2.33%-3.23%) | 69.37% (68.08%-70.65%) | 3.67% (3.14%-4.19%) |
| 8 | 2005 | 8 | 73.08% (72%-74.16%) | 6.04% (5.46%-6.61%) | 70.15% (69.01%-71.3%) | 4.31% (3.8%-4.81%) | 74.28% (73.16%-75.41%) | 5.99% (5.38%-6.6%) |
| 8 | 2005 | 7 | 73.73% (72.64%-74.81%) | 6.38% (5.78%-6.99%) | 70.63% (69.5%-71.76%) | 4.7% (4.17%-5.23%) | 74.26% (73.15%-75.38%) | 6.1% (5.49%-6.71%) |
| 8 | 2005 | 6 | 74.07% (72.97%-75.16%) | 6.05% (5.46%-6.65%) | 71.82% (70.67%-72.96%) | 4.91% (4.36%-5.46%) | 75.31% (74.18%-76.43%) | 6.09% (5.47%-6.72%) |
| 8 | 2005 | 5 | 73.92% (72.8%-75.04%) | 6.29% (5.67%-6.91%) | 71.84% (70.69%-73%) | 5.09% (4.52%-5.65%) | 75.67% (74.54%-76.8%) | 5.86% (5.24%-6.48%) |
| 8 | 2005 | 4 | 75.86% (74.79%-76.94%) | 5.99% (5.39%-6.58%) | 71.99% (70.84%-73.15%) | 5.26% (4.68%-5.83%) | 76.03% (74.9%-77.16%) | 6.52% (5.87%-7.18%) |
| 8 | 2005 | 3 | 74.94% (73.92%-75.97%) | 6.02% (5.45%-6.58%) | 72.63% (71.55%-73.71%) | 5.39% (4.84%-5.93%) | 76.51% (75.46%-77.56%) | 6.7% (6.08%-7.32%) |
| 8 | 2005 | 2 | 74.32% (73.26%-75.38%) | 6% (5.42%-6.58%) | 74.01% (72.92%-75.11%) | 5.28% (4.72%-5.84%) | 77.02% (75.94%-78.1%) | 6.3% (5.67%-6.92%) |
| 8 | 2005 | 1 | 75.05% (74.07%-76.03%) | 5.69% (5.17%-6.22%) | 72.54% (71.47%-73.61%) | 5.17% (4.64%-5.7%) | 75.88% (74.83%-76.93%) | 6.57% (5.96%-7.18%) |
| 8 | 2004 | 12 | 74.74% (73.77%-75.72%) | 5.88% (5.35%-6.41%) | 72.36% (71.3%-73.42%) | 4.65% (4.15%-5.15%) | 75.01% (73.95%-76.06%) | 5.92% (5.35%-6.5%) |
| 8 | 2004 | 11 | 74.4% (73.41%-75.4%) | 5.93% (5.39%-6.47%) | 72.09% (71.01%-73.16%) | 4.61% (4.1%-5.11%) | 75.51% (74.45%-76.57%) | 5.69% (5.12%-6.26%) |
| 8 | 2004 | 10 | 73.5% (72.45%-74.56%) | 5.92% (5.36%-6.49%) | 73.17% (72.07%-74.27%) | 4.66% (4.14%-5.18%) | 76.86% (75.79%-77.94%) | 6% (5.4%-6.61%) |
| 8 | 2004 | 9 | 71.85% (70.51%-73.19%) | 5.27% (4.61%-5.94%) | 72.57% (71.36%-73.79%) | 5.58% (4.96%-6.21%) | 75.27% (74.07%-76.48%) | 7.22% (6.49%-7.94%) |
| 9 | 2004 | 8 | 76.96% (75.9%-78.02%) | 6.9% (6.26%-7.53%) | 75.35% (74.24%-76.46%) | 7.05% (6.39%-7.71%) | 77.82% (76.71%-78.94%) | 8.16% (7.43%-8.89%) |
| 9 | 2004 | 7 | 76.17% (75.08%-77.26%) | 7.06% (6.4%-7.72%) | 75.2% (74.07%-76.33%) | 7.25% (6.57%-7.93%) | 77.56% (76.42%-78.7%) | 8.41% (7.65%-9.17%) |
| 9 | 2004 | 6 | 76.64% (75.52%-77.75%) | 7.85% (7.14%-8.56%) | 75.77% (74.64%-76.91%) | 7.14% (6.46%-7.83%) | 78.16% (77.02%-79.3%) | 8.03% (7.28%-8.78%) |
| 9 | 2004 | 5 | 77.27% (76.15%-78.38%) | 7.94% (7.22%-8.66%) | 76.25% (75.1%-77.4%) | 7.42% (6.71%-8.13%) | 77.82% (76.65%-78.98%) | 8.45% (7.67%-9.23%) |
| 9 | 2004 | 4 | 77.74% (76.64%-78.85%) | 9.05% (8.28%-9.81%) | 77.8% (76.7%-78.9%) | 7.3% (6.61%-7.99%) | 80.12% (79.01%-81.23%) | 8.66% (7.88%-9.44%) |
| 9 | 2004 | 3 | 78.6% (77.58%-79.63%) | 8.45% (7.76%-9.15%) | 77.27% (76.22%-78.32%) | 7.43% (6.77%-8.09%) | 78.7% (77.63%-79.77%) | 8.8% (8.05%-9.54%) |

| | | | | | | | | |
|---|---|---|---|---|---|---|---|---|
| 9 | 2004 | 2 | 77.13% (76.07%-78.19%) | 7.63% (6.97%-8.3%) | 76.56% (75.46%-77.65%) | 7.27% (6.6%-7.95%) | 78.64% (77.53%-79.75%) | 8.45% (7.7%-9.2%) |
| 9 | 2004 | 1 | 77.58% (76.52%-78.64%) | 7.52% (6.85%-8.19%) | 77.67% (76.67%-78.68%) | 6.53% (5.93%-7.12%) | 79.03% (78.01%-80.06%) | 7.91% (7.23%-8.59%) |
| 9 | 2003 | 12 | 76.55% (75.44%-77.66%) | 7.95% (7.24%-8.65%) | 76.37% (75.36%-77.38%) | 6.9% (6.3%-7.5%) | 78.66% (77.64%-79.67%) | 8.21% (7.53%-8.89%) |
| 9 | 2003 | 11 | 75.64% (74.47%-76.81%) | 6.75% (6.07%-7.44%) | 76.75% (75.73%-77.77%) | 7.29% (6.66%-7.92%) | 78.57% (77.53%-79.6%) | 8.08% (7.39%-8.77%) |
| 9 | 2003 | 10 | 76.45% (75.2%-77.69%) | 6.83% (6.09%-7.57%) | 75.28% (74.18%-76.37%) | 6.67% (6.03%-7.3%) | 77.79% (76.69%-78.89%) | 8.27% (7.54%-9%) |
| 9 | 2003 | 9 | 76.18% (74.59%-77.77%) | 6.57% (5.65%-7.5%) | 75.12% (73.75%-76.48%) | 6.75% (5.96%-7.55%) | 76.75% (75.36%-78.13%) | 7.66% (6.78%-8.53%) |
| 10 | 2003 | 8 | 79.25% (78.06%-80.44%) | 9.38% (8.52%-10.23%) | 79.44% (78.27%-80.61%) | 8.68% (7.87%-9.49%) | 80.92% (79.75%-82.09%) | 9.91% (9.02%-10.8%) |
| 10 | 2003 | 7 | 79.69% (78.5%-80.87%) | 10.58% (9.68%-11.49%) | 79.98% (78.8%-81.16%) | 9.37% (8.51%-10.23%) | 80.46% (79.26%-81.66%) | 10.44% (9.51%-11.36%) |
| 10 | 2003 | 6 | 79.53% (78.32%-80.75%) | 9.84% (8.95%-10.74%) | 79.5% (78.28%-80.73%) | 9.94% (9.03%-10.84%) | 80.91% (79.68%-82.14%) | 11.45% (10.46%-12.45%) |
| 10 | 2003 | 5 | 80.43% (79.22%-81.64%) | 10.87% (9.92%-11.82%) | 80.21% (78.99%-81.42%) | 10.15% (9.23%-11.07%) | 80.2% (78.96%-81.45%) | 11% (10.02%-11.98%) |
| 10 | 2003 | 4 | 80.33% (79.14%-81.53%) | 10.73% (9.8%-11.65%) | 80.46% (79.25%-81.66%) | 10.61% (9.68%-11.55%) | 81.32% (80.1%-82.53%) | 11.74% (10.74%-12.75%) |
| 10 | 2003 | 3 | 80.26% (79.13%-81.39%) | 10.36% (9.5%-11.23%) | 81.41% (80.29%-82.53%) | 10.42% (9.54%-11.3%) | 82.25% (81.12%-83.38%) | 11.51% (10.56%-12.45%) |
| 10 | 2003 | 2 | 80.07% (78.9%-81.24%) | 10.05% (9.17%-10.93%) | 79.19% (78.02%-80.36%) | 8.84% (8.03%-9.66%) | 80.12% (78.93%-81.31%) | 9.85% (8.96%-10.74%) |
| 10 | 2003 | 1 | 80.17% (79.04%-81.29%) | 9.82% (8.99%-10.66%) | 79.38% (78.2%-80.57%) | 9.39% (8.54%-10.25%) | 80.48% (79.29%-81.67%) | 10.05% (9.15%-10.96%) |
| 10 | 2002 | 12 | 79.21% (78.06%-80.37%) | 9.19% (8.36%-10.01%) | 80.13% (78.93%-81.33%) | 9.42% (8.54%-10.3%) | 81.13% (79.91%-82.34%) | 10.42% (9.47%-11.37%) |
| 10 | 2002 | 11 | 78.73% (77.52%-79.94%) | 8.84% (8%-9.67%) | 78.08% (76.78%-79.37%) | 8.65% (7.77%-9.53%) | 80.12% (78.83%-81.41%) | 9.68% (8.73%-10.64%) |
| 10 | 2002 | 10 | 78.94% (77.61%-80.27%) | 9.23% (8.29%-10.18%) | 79.75% (78.39%-81.1%) | 9.08% (8.11%-10.05%) | 80.49% (79.12%-81.87%) | 9.86% (8.83%-10.9%) |
| 10 | 2002 | 9 | 75.85% (74.06%-77.63%) | 8.08% (6.95%-9.22%) | 79.44% (77.72%-81.16%) | 8.27% (7.1%-9.44%) | 79.28% (77.5%-81.07%) | 9.53% (8.23%-10.82%) |
| 11 | 2002 | 8 | 81.42% (80.27%-82.57%) | 10.72% (9.8%-11.63%) | 80.12% (78.87%-81.36%) | 10.88% (9.91%-11.85%) | 80.23% (78.95%-81.51%) | 11.56% (10.53%-12.59%) |
| 11 | 2002 | 7 | 82.13% (80.99%-83.28%) | 12.57% (11.58%-13.56%) | 81.16% (79.94%-82.38%) | 11.53% (10.53%- | 82.26% (81.02%-83.49%) | 13.15% (12.06%- |

| | | | | | | | | |
|---|---|---|---|---|---|---|---|---|
| | | | | | 12.53%) | | 14.24%) |
| 11 | 2002 | 6 | 80.61% (79.39%-81.83%) | 11.63% (10.64%-12.62%) | 79.98% (78.7%-81.26%) | 11.2% (10.19%-12.21%) | 80.97% (79.67%-82.26%) | 12.33% (11.24%-13.42%) |
| 11 | 2002 | 5 | 81.79% (80.6%-82.98%) | 12.12% (11.12%-13.13%) | 81% (79.74%-82.27%) | 12.18% (11.12%-13.23%) | 82.05% (80.77%-83.32%) | 12.95% (11.83%-14.06%) |
| 11 | 2002 | 4 | 83.16% (82.01%-84.31%) | 12.25% (11.24%-13.26%) | 81.08% (79.83%-82.33%) | 12.47% (11.42%-13.52%) | 81.82% (80.55%-83.1%) | 13.09% (11.98%-14.21%) |
| 11 | 2002 | 3 | 82.33% (81.21%-83.44%) | 11.96% (11.01%-12.91%) | 80.9% (79.72%-82.09%) | 11.42% (10.46%-12.37%) | 81.71% (80.51%-82.91%) | 13.09% (12.04%-14.13%) |
| 11 | 2002 | 2 | 82.43% (81.28%-83.57%) | 11.49% (10.53%-12.45%) | 80.77% (79.54%-82%) | 11.35% (10.36%-12.34%) | 81.25% (79.99%-82.5%) | 11.81% (10.77%-12.85%) |
| 11 | 2002 | 1 | 82.48% (81.34%-83.62%) | 11.56% (10.61%-12.52%) | 81.66% (80.49%-82.83%) | 11.54% (10.58%-12.5%) | 83.02% (81.85%-84.19%) | 12.3% (11.27%-13.32%) |
| 11 | 2001 | 12 | 80.76% (79.55%-81.97%) | 11.59% (10.61%-12.58%) | 79.72% (78.51%-80.94%) | 10.62% (9.68%-11.55%) | 80.56% (79.32%-81.79%) | 12.2% (11.18%-13.22%) |
| 11 | 2001 | 11 | 81.71% (80.47%-82.96%) | 11.26% (10.24%-12.28%) | 79.78% (78.52%-81.03%) | 9.97% (9.03%-10.91%) | 80.97% (79.7%-82.24%) | 10.76% (9.76%-11.77%) |
| 11 | 2001 | 10 | 79.52% (78.15%-80.89%) | 10.33% (9.3%-11.37%) | 80.09% (78.71%-81.47%) | 10.1% (9.06%-11.14%) | 81.27% (79.88%-82.66%) | 11.28% (10.15%-12.41%) |
| 11 | 2001 | 9 | 82.04% (80.36%-83.72%) | 10.47% (9.13%-11.81%) | 77.73% (75.9%-79.55%) | 9.91% (8.6%-11.22%) | 78.67% (76.82%-80.52%) | 10.59% (9.19%-11.98%) |
| 12 | 2001 | 8 | 82.74% (81.47%-84.01%) | 13.08% (11.95%-14.21%) | 84.28% (83.02%-85.54%) | 12.98% (11.82%-14.14%) | 85.29% (83.99%-86.59%) | 13.98% (12.71%-15.25%) |
| 12 | 2001 | 7 | 83.41% (82.15%-84.68%) | 12.45% (11.32%-13.57%) | 85.88% (84.68%-87.09%) | 14.8% (13.57%-16.04%) | 86.57% (85.32%-87.82%) | 16.22% (14.87%-17.57%) |
| 12 | 2001 | 6 | 81.15% (79.81%-82.49%) | 13.8% (12.62%-14.98%) | 84.01% (82.69%-85.32%) | 14.65% (13.38%-15.92%) | 85.54% (84.19%-86.89%) | 15.68% (14.29%-17.08%) |
| 12 | 2001 | 5 | 82.53% (81.24%-83.83%) | 13.35% (12.19%-14.51%) | 84.98% (83.71%-86.25%) | 14.5% (13.25%-15.75%) | 85.36% (84.03%-86.69%) | 15.71% (14.34%-17.08%) |
| 12 | 2001 | 4 | 82.42% (81.13%-83.72%) | 12.27% (11.15%-13.39%) | 85.47% (84.21%-86.72%) | 14.53% (13.28%-15.79%) | 87.25% (85.99%-88.52%) | 16.01% (14.62%-17.4%) |
| 12 | 2001 | 3 | 82.49% (81.23%-83.74%) | 13.31% (12.19%-14.44%) | 85.45% (84.25%-86.65%) | 14.1% (12.92%-15.28%) | 86.81% (85.59%-88.03%) | 15.02% (13.73%- |

| | | | | | | | | |
|---|---|---|---|---|---|---|---|---|
| | | | | | | | | 16.31%) |
| 12 | 2001 | 2 | 82.76% (81.49%-84.03%) | 11.75% (10.67%-12.83%) | 83.68% (82.39%-84.96%) | 14.77% (13.54%-16.01%) | 85.7% (84.4%-87%) | 15.23% (13.9%-16.56%) |
| 12 | 2001 | 1 | 82.51% (81.31%-83.7%) | 13.1% (12.03%-14.16%) | 86.01% (84.8%-87.21%) | 13.9% (12.69%-15.1%) | 85.93% (84.65%-87.21%) | 14.88% (13.57%-16.2%) |

**Supplemental Table S7. The influence of COVID-19 on myopia progression among school-aged children**

| | Normal (6-month interval) | | COVID-19 (6-month interval) | | P value |
|---|---|---|---|---|---|
| **Characteristics** | **Count** | **Mean myopia progression(95% CI)** | **Count** | **Mean myopia progression (95% CI)** | |
| **Total** | 547,475 | -0.263 (-0.262, -0.264) | 575,597 | -0.390 (-0.389, -0.391) | < 0.001 |
| **Gender** | | | | | |
| Male | 299,857 | -0.266 (-0.264, -0.268) | 314,432 | -0.385(-0.383, -0.387) | < 0.001 |
| Female | 247,618 | -0.259 (-0.257, -0.261) | 261,165 | -0.396(-0.394, -0.398) | < 0.001 |
| **Educational level** | | | | | |
| Grade group I | 325,756 | -0.259 (-0.258, -0.261) | 349,152 | -0.442 (-0.440, -0.443) | < 0.001 |
| Grade group II | 151,446 | -0.277 (-0.274, -0.280) | 155,708 | -0.327 (-0.324, -0.329) | < 0.001 |
| **Educational system** | | | | | |
| Non-Key school | 106,107 | -0.263 (-0.261, -0.264) | 118,971 | -0.395 (-0.394, -0.397) | < 0.001 |
| Key school | 441,368 | -0.264 (-0.261, -0.267) | 456,626 | -0.369 (-0.366, -0.372) | < 0.001 |
| **Region of habitation** | | | | | |
| Rural | 426,584 | -0.255 (-0.252, -0.258) | 444,595 | -0.392 (-0.389, -0.396) | < 0.001 |
| Urban | 90,632 | -0.263 (-0.262, -0.265) | 89,517 | -0.388 (-0.387, -0.390) | < 0.001 |

**Supplemental Table S8. The impact of COVID-19 on myopia progression among school-aged children categorized by grade**

| | | | Normal (6-month interval) | | COVID-19 (6-month interval) | | P value |
|---|---|---|---|---|---|---|---|
| **Grade group** | **Characteristics** | **Grade** | **Count** | **Mean myopia progression (95%CI)** | **Count** | **Mean myopia progression (95%CI)** | |
| **Grade Group I** | Elementary School | 1 | 54,648 | -0.225 (-0.222, -0.229) | 60,977 | -0.418 (-0.414, -0.422) | < 0.001 |
| | | 2 | 53,537 | -0.224 (-0.221, -0.228) | 58,965 | -0.441(-0.437, -0.445) | < 0.001 |
| | | 3 | 53,996 | -0.240 (-0.236, -0.244) | 57,941 | -0.463(-0.459, -0.467) | < 0.001 |
| | | 4 | 55,713 | -0.263 (-0.260, -0.267) | 59,030 | -0.461(-0.457, -0.465) | < 0.001 |
| | | 5 | 56,615 | -0293 (-0.289, -0.297) | 58,917 | -0.464(-0.460, -0.469) | < 0.001 |
| | | 6 | 51,247 | -0.311 (-0.306, -0.316) | 53,322 | -0.40 (-0.396, -0.404) | < 0.001 |
| **Grade Group II** | Junior high school | 7 | 57,311 | -0.272 (-0.268, -0.275) | 58,656 | -0.352 (-0.348, -0.356) | < 0.001 |
| | | 8 | 54,135 | -0.268 (-0.264, -0.272) | 54,587 | -0.323 (-0.319, -0.327) | < 0.001 |
| | | 9 | 40,000 | -0.297 (-0.290, -0.303) | 42,465 | -0.297 (-0.292, -0.302) | 0.96 |
| | Senior high school | 10 | 39,773 | -0.240 (-0.235, -0.245) | 40,322 | -0.268(-0.263, -0.273) | < 0.001 |
| | | 11 | 30,500 | -0.260 (-0.254, -0.267) | 30,415 | -0.281 (-0.275, -0.287) | < 0.001 |

**Supplemental Table S9. The impact of COVID-19 on incidence of myopia among school-aged children categorized by grade**

| Characteristics | Grade | Normal (6-month interval) | | COVID-19 (6-month interval) | | Odds Ratio (95% CI) | P value |
|---|---|---|---|---|---|---|---|
| | | Count | Incidence rate (95% CI) | Count | Incidence rate (95% CI) | | |
| Elementary School | 1 | 52,049 | 4.66% (4.48%-4.84%) | 49,622 | 7.64% (7.40%-7.87%) | 1.69 (1.60-1.78) | < 0.001 |
| | 2 | 43,438 | 6.21% (5.98%-6.43%) | 40,742 | 9.85% (9.56%-10.13%) | 1.65 (1.57-1.74) | < 0.001 |
| | 3 | 34,587 | 8.54% (8.25%-8.84%) | 31,632 | 12.05% (11.69%-12.41%) | 1.47 (1.39-1.54) | < 0.001 |
| | 4 | 27,220 | 9.89% (9.54%-10.24%) | 24,528 | 12.34% (11.93%-12.75%) | 1.28 (1.21-1.36) | < 0.001 |
| | 5 | 19,798 | 11.88% (11.43%-12.33%) | 17,446 | 13.66% (13.15%-14.17%) | 1.17 (1.10-1.25) | < 0.001 |
| | 6 | 13,541 | 13.82% (13.24%-14.41%) | 11,669 | 11.10% (10.53%-11.67%) | 0.78 (0.72-0.84) | < 0.001 |
| Junior high school | 7 | 10,371 | 12.15% (11.52%-12.78%) | 9,111 | 8.34% (7.77%-8.91%) | 0.66 (0.60-0.72) | < 0.001 |
| | 8 | 7,880 | 12.09% (11.37%-12.81%) | 6,927 | 7.10% (6.50%-7.71%) | 0.56 (0.50-0.62) | < 0.001 |
| | 9 | 4,813 | 16.77% (15.71%-17.82%) | 4,006 | 3.52% (2.95%-4.09%) | 0.18 (0.15-0.22) | < 0.001 |
| Senior high school | 10 | 4,410 | 9.95% (9.07%-10.84%) | 3,971 | 3.05% (2.51%-3.58%) | 0.28 (0.23-0.35) | < 0.001 |
| | 11 | 2,297 | 12.7% (11.35%-14.08%) | 2,005 | 3.84% (3.00%-4.68%) | 0.27 (0.21-0.36) | < 0.001 |

**Supplemental Table S10. The impact of COVID-19 on incidence of high myopia among school-aged children categorized by grade**

| Characteristics | Grade | Normal (6-month interval) | | COVID-19 (6-month interval) | | Odds Ratio (95% CI) | P value |
|---|---|---|---|---|---|---|---|
| | | Count | Incidence rate (95% CI) | Count | Incidence rate (95% CI) | | |
| Elementary School | 1 | 58,658 | 0.063% (0.043%-0.083%) | 58,621 | 0.061% (0.041%-0.081%) | 0.97 (0.60-1.58) | 1.0 |
| | 2 | 55,552 | 0.11% (0.084%-0.14%) | 55,490 | 0.16% (0.13%-0.19%) | 1.44 (1.03-2.02) | 0.028 |
| | 3 | 54,341 | 0.23% (0.19%-0.27%) | 54,214 | 0.39% (0.33%-0.44%) | 1.65 (1.32-2.08) | < 0.001 |
| | 4 | 55,753 | 0.45% (0.39%-0.51%) | 55,501 | 0.63% (0.57%-0.70%) | 1.41 (1.91-1.66) | < 0.001 |

| | | | | | | | |
|---|---|---|---|---|---|---|---|
| | 5 | 56,440 | 0.77% (0.69%-0.84%) | 56,008 | 1.17% (1.08%-1.26%) | 1.54 (1.36-1.74) | < 0.001 |
| | 6 | 51,554 | 1.32% (1.22%-1.42%) | 50,873 | 1.41% (1.31%-1.51%) | 1.07 (0.96-1.19) | 0.22 |
| Junior high school | 7 | 56,667 | 1.64% (1.54%-1.75%) | 55,736 | 1.59% (1.48%-1.69%) | 0.97 (0.88-1.06) | 0.48 |
| | 8 | 52,790 | 2.19% (2.07%-2.32%) | 51,632 | 1.61% (1.51%-1.72%) | 0.73 (0.67-0.80) | < 0.001 |
| | 9 | 40,745 | 3.13% (2.96%-3.30%) | 39,469 | 1.48% (1.36%-1.59%) | 0.46 (0.42-0.51) | < 0.001 |
| Senior high school | 10 | 38,117 | 2.97% (2.80%-3.14%) | 36,985 | 1.38% (1.26%-1.50%) | 046 (0.41-0.51) | < 0.001 |
| | 11 | 28,605 | 3.87% (3.65%-4.09%) | 27,498 | 1.13% (1.01%-1.26%) | 0.28 (0.25-0.32) | < 0.001 |

#### References:

1. You QS, Wu LJ, Duan JL, et al. Prevalence of myopia in school children in greater Beijing: the Beijing Childhood Eye Study. *Acta ophthalmologica* 2014; **92**(5): e398-e406.
2. You QS, Wu LJ, Duan JL, et al. Factors associated with myopia in school children in China: the Beijing childhood eye study. *PloS one* 2012; **7**(12): e52668.
3. Wu LJ, You QS, Duan JL, et al. Prevalence and associated factors of myopia in high-school students in Beijing. *PLoS One* 2015; **10**(3): e0120764.
4. Zhao J, Pan X, Sui R, Munoz SR, Sperduto RD, Ellwein LB. Refractive error study in children: results from Shunyi District, China. *American journal of ophthalmology* 2000; **129**(4): 427-35.
5. Lyu Y, Zhang H, Gong Y, et al. Prevalence of and factors associated with myopia in primary school students in the Chaoyang District of Beijing, China. *Japanese journal of ophthalmology* 2015; **59**(6): 421-9.
6. Ye S, Liu S, Li W, Wang Q, Xi W, Zhang X. Associations between anthropometric indicators and both refraction and ocular biometrics in a cross-sectional study of Chinese schoolchildren. *BMJ open* 2019; **9**(5): e027212.
7. He M, Huang W, Zheng Y, Huang L, Ellwein LB. Refractive error and visual impairment in school children in rural southern China. *Ophthalmology* 2007; **114**(2): 374-82. e1.
8. He M, Zeng J, Liu Y, Xu J, Pokharel GP, Ellwein LB. Refractive error and visual impairment in urban children in southern China. *Investigative ophthalmology & visual science* 2004; **45**(3): 793-9.
9. Guo K, Yang DY, Wang Y, et al. Prevalence of myopia in schoolchildren in Ejina: the Gobi Desert children eye study. *Investigative ophthalmology & visual science* 2015; **56**(3): 1769-74.
10. Wu JF, Bi HS, Wang SM, et al. Refractive error, visual acuity and causes of vision loss in children in Shandong, China. The Shandong Children Eye Study. *PLoS One* 2013; **8**(12): e82763.
11. Li Z, Xu K, Wu S, et al. Population‐based survey of refractive error among school‐aged children in rural northern China: the Heilongjiang eye study. *Clinical & experimental ophthalmology* 2014; **42**(4): 379-84.
12. Pi L-H, Chen L, Liu Q, et al. Refractive status and prevalence of refractive errors in suburban school-age children. *International journal of medical sciences* 2010; **7**(6): 342.
13. MORGAN A, YOUNG R, NARANKHAND B, CHEN S, COTTRIALL C, HOSKING S. Prevalence rate of myopia in schoolchildren in rural Mongolia. *Optometry and vision science* 2006; **83**(1): 53-6.
14. Congdon N, Wang Y, Song Y, et al. Visual disability, visual function, and myopia among rural Chinese secondary school children: the Xichang Pediatric Refractive Error Study (X-PRES)—report 1. *Investigative ophthalmology & visual science*

2008; **49**(7): 2888-94.
15. Fan DS, Lam DS, Lam RF, et al. Prevalence, incidence, and progression of myopia of school children in Hong Kong. *Investigative ophthalmology & visual science* 2004; **45**(4): 1071-5.
16. Lam CSY, Lam CH, Cheng SCK, Chan LYL. Prevalence of myopia among Hong Kong Chinese schoolchildren: changes over two decades. *Ophthalmic and Physiological Optics* 2012; **32**(1): 17-24.
17. Lam CSY, Goldschmidt E, Edwards MH. Prevalence of myopia in local and international schools in Hong Kong. *Optometry and vision science* 2004; **81**(5): 317-22.
18. LL L, YF S, CK H, CJ C, LA L, PT H. Epidemiologic study of the prevalence and severity of myopia among schoolchildren in Taiwan in 2000. *Journal of the Formosan Medical Association = Taiwan yi zhi* 2001; **100**(10): 684-91.
19. Lin LL-K, Shih Y-F, Hsiao CK, Chen C. Prevalence of myopia in Taiwanese schoolchildren: 1983 to 2000. *Annals Academy of Medicine Singapore* 2004; **33**(1): 27-33.
20. Lai Y-H, Hsu H-T, Wang H-Z, Chang S-J, Wu W-C. The visual status of children ages 3 to 6 years in the vision screening program in Taiwan. *Journal of American Association for Pediatric Ophthalmology and Strabismus* 2009; **13**(1): 58-62.
21. Guo Y, Lin H, Lin L, Cheng C. Self-reported myopia in Taiwan: 2005 Taiwan national health interview survey. *Eye* 2012; **26**(5): 684.
22. Lee JH, Jee D, Kwon J-W, Lee WK. Prevalence and risk factors for myopia in a rural Korean population. *Investigative ophthalmology & visual science* 2013; **54**(8): 5466-71.
23. Rim TH, Kim S-H, Lim KH, Choi M, Kim HY, Baek S-H. Refractive errors in koreans: the Korea national health and nutrition examination survey 2008-2012. *Korean Journal of Ophthalmology* 2016; **30**(3): 214-24.
24. Lim HT, Yoon JS, Hwang S-S, Lee SY. Prevalence and associated sociodemographic factors of myopia in Korean children: the 2005 third Korea National Health and Nutrition Examination Survey (KNHANES III). *Japanese journal of ophthalmology* 2012; **56**(1): 76-81.
25. Jung S-K, Lee JH, Kakizaki H, Jee D. Prevalence of myopia and its association with body stature and educational level in 19-year-old male conscripts in Seoul, South Korea. *Investigative ophthalmology & visual science* 2012; **53**(9): 5579-83.
26. Quek TP, Chua CG, Chong CS, et al. Prevalence of refractive errors in teenage high school students in Singapore. *Ophthalmic and Physiological Optics* 2004; **24**(1): 47-55.
27. Saw S-M, Carkeet A, Chia K-S, Stone RA, Tan DT. Component dependent risk factors for ocular parameters in Singapore Chinese children. *Ophthalmology* 2002; **109**(11): 2065-71.
28. Rose KA, Morgan IG, Smith W, Burlutsky G, Mitchell P, Saw S-M. Myopia, lifestyle, and schooling in students of Chinese ethnicity in Singapore and Sydney. *Archives of ophthalmology* 2008; **126**(4): 527-30.
29. Matsumura H, Hirai H. Prevalence of myopia and refractive changes in students from 3 to 17 years of age. *Survey of ophthalmology* 1999; **44**: S109-S15.
30. Yotsukura E, Torii H, Inokuchi M, et al. Current Prevalence of Myopia and Association of Myopia With Environmental Factors Among Schoolchildren in Japan. *JAMA ophthalmology* 2019.
